# Evaluation of polygenic risk scores and ambient air pollutants for lung cancer risk stratification in a lung cancer screening cohort

**DOI:** 10.64898/2026.07.14.26358054

**Authors:** Lianne Trap, Ramazan Büyükcelik, Noa Antonissen, Grigory A. Sidorenkov, Rikje Ruiter, Jolande van Heemst, Bahar Sedaghati-Khayat, Bernard S. Stikker, Daphne W. Dumoulin, Hester A. Gietema, Marjolein A. Heuvelmans, Firdaus A.A. Mohamed Hoesein, Pim A. de Jong, André G. Uitterlinden, Guy Brusselle, Colin Jacobs, Joachim G.J.V. Aerts, Roel C.H. Vermeulen, Geertruida H. De Bock, Harry J.M. Groen, Rozemarijn Vliegenthart, George S. Downward, Ralph Stadhouders, Jeroen van Rooij, the NELSON-POP consortium

## Abstract

**Background:** Randomized controlled trials have shown that computed tomographic (CT) screening reduces lung cancer mortality. Improved identification of at-risk groups, by leveraging non-smoking risk factors, could help refine screening selection.

**Aim:** To evaluate polygenic risk scores (PRSs) and ambient air pollution (AAP) exposure for risk stratification in the NELSON lung cancer screening cohort.

**Methods:** Two PRSs (PRS-McKay/PRS-Byun) and several AAPs (including nitrogen dioxide, ozone, and particulate matter [PM]) were assessed in the NELSON lung cancer screening trial (N=7,364). PRSs were validated in the Rotterdam Study (N=11,493). Associations with lung cancer, mortality, screening results, and discriminative ability to distinguish lung cancer were evaluated.

**Results:** PRS-McKay and PRS-Byun were associated with lung cancer (odds ratio [OR] per SD [95%CI]: 1.22 [1.08-1.37] and 1.28 [1.13-1.44], respectively) and lung cancer-specific mortality (OR [95%CI]: 1.24 [1.05-1.47], for both), but not with non-lung cancer mortality (OR [95%CI]: 1.01 [0.94-1.10] and 1.03 [0.95-1.12], respectively). Exposure to PM_2.5_ was associated with lung cancer (OR [95%CI]: 1.11 [1.01-1.22]). PM constituents were associated with adenocarcinoma, particularly PM_10_ (OR [95%CI]: 1.16 [1.01-1.32]) and ultra-fine particles (OR [95%CI]: 1.16 [1.04-1.30]). PRS and AAP added modestly to the discriminative ability for lung cancer on top of pack-years, age, and sex (area under the curve [95%CI]: 0.659 [0.624-0.695] vs. 0.643 [0.608-0.679]).

**Conclusions:** PRSs and exposure to PM were associated with lung cancer in a high-risk screening population. The primary potential of PRSs may reside in refining lung cancer screening selection toward individuals at higher risk of dying from lung cancer specifically.

## INTRODUCTION

Lung cancer is the primary cause of cancer mortality and morbidity globally, with nearly 2.5 million new cases and over 1.8 million deaths in 2022 alone.^1^ Low-dose computed tomographic (CT) screening has been shown to be effective in reducing lung cancer mortality by at least 20% in high-risk individuals.^2,3^

The primary and historically used eligibility criteria for lung cancer screening are age and smoking history.^4^ Multivariable risk stratification models enhance the accuracy and predictive ability of selection based on age and smoking history alone,^5,6^ and are recommended for participant selection by the European Union.^7^ Individual variation in genetic risk, beyond family history, and exposure to ambient air pollution have been consistently linked to lung cancer risk.^8–21^ However, current lung cancer risk stratification models and screening eligibility criteria do not incorporate either factor.

Polygenic risk scores (PRSs) estimate an individual’s lifetime genetic risk of disease. Notably, individuals in the highest PRS percentiles can have a similar disease risk to those with pathogenic monogenic variants.^22^ In breast cancer screening, genetic stratification including a PRS is already incorporated to personalize screening recommendations (model: BOADICEA, tool: CanRisk), which has improved risk prediction and informed screening initiation age and frequency.^23^

Ambient air pollution (AAP) is considered carcinogenic to humans (IARC Group 1)^24^ and ambient particulate matter <2.5 μm in diameter (PM_2.5_) is the second-leading risk factor for lung cancer mortality worldwide, following tobacco smoking.^25,26^ Additionally, there is increasing interest in the role of ultra-fine-particles (UFP; particulate matter <0.1 μm), although this remains largely understudied. The impact of AAP on lung cancer risk varies by histological subtype, with the strongest associations observed in adenocarcinoma,^13,19,27^ now the most common subtype worldwide.^28^

Although PRS and AAP have been studied in the general population and in case-control settings, their associations with lung cancer outcomes in screening participants remain underexplored. We therefore evaluated the performance of PRS and AAP in the Nederlands– Leuvens Longkanker Screenings Onderzoek (NELSON) screening arm. Specifically, we examined whether PRSs and AAP exposures are associated with lung cancer incidence, mortality, and screening results, and assessed their discriminative ability to distinguish lung cancer. The PRSs were additionally validated in an independent prospective cohort (The Rotterdam Study).

## MATERIALS AND METHODS

### Study overview

We examined the association and discriminative ability of two PRSs and five AAP constituents (nitrogen dioxide [NO_2_], ozone [O_3_], and PM_10_, PM_2.5_, and ultra-fine particles [UFP]) with lung cancer risk in the NELSON cohort. The discriminative ability of the PRSs was validated in the Rotterdam Study (RS), a cohort of inhabitants from a single suburb in Rotterdam, The Netherlands. An overview of the variables studied in each cohort, including exposures, outcomes, and the overall analytical framework is shown in **Figure 1**.

**Figure 1.**
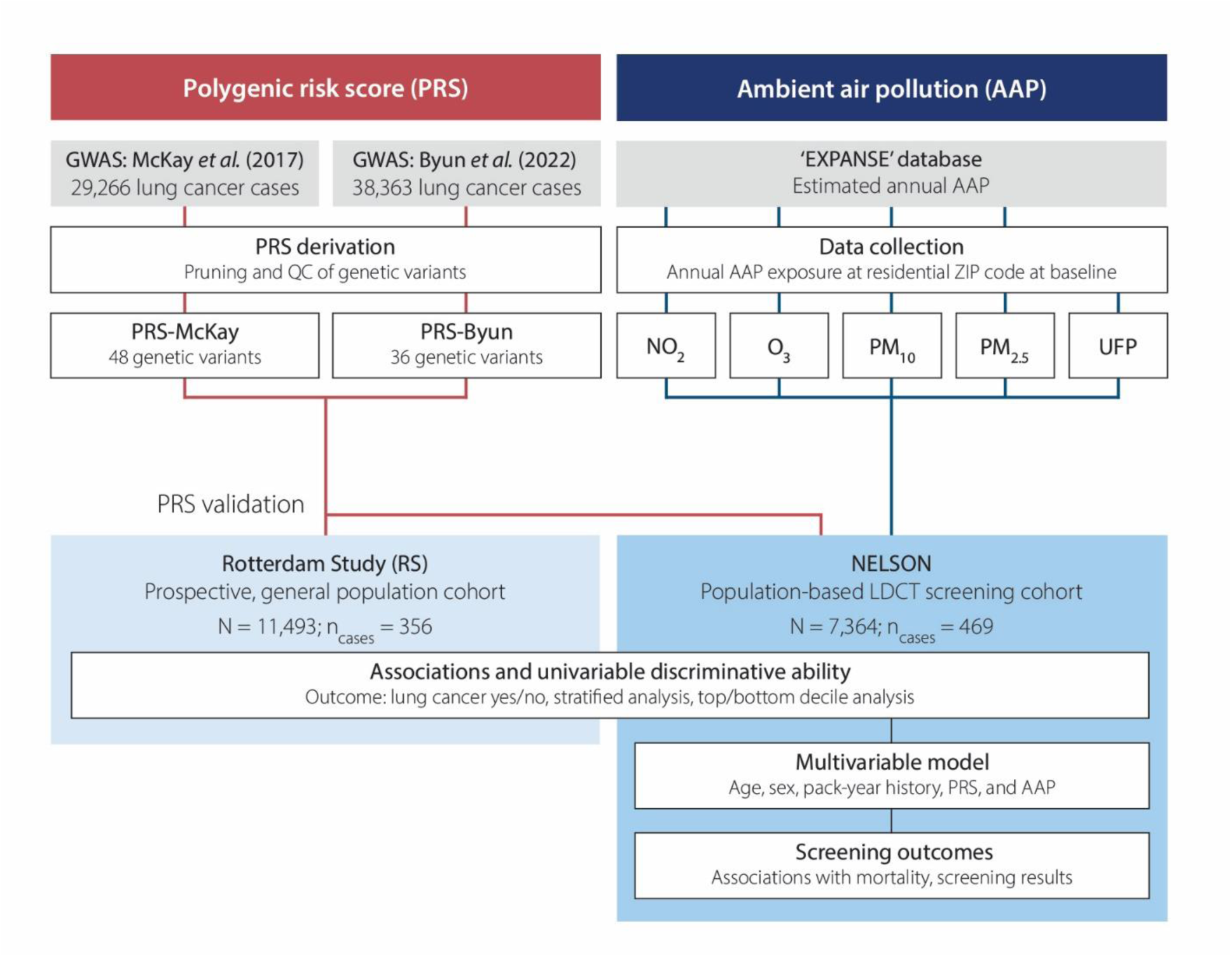
Study overview. Schematic representation of the study, including the derivation of the PRSs, AAP data collection, study cohorts, outcomes assessed, and the analyses performed. LDCT: low-dose computed tomography. NO_2_: nitrogen dioxide, O_3_: ozone, PM_10_: particulate matter <10 μm in diameter, PM_2.5_: particulate matter <2.5 μm in diameter, UFP: ultra-fine particles; <0.1 μm in diameter.

### Study cohorts: the Rotterdam Study (RS)

The RS is a Dutch, prospective, mid-late life cohort, which primarily investigates diseases of old age.^29^ See **Supplementary Methods** for additional details on cohort description and genetic data. In this study we used data from the genotyped proportion of RS (n_RS-I_=6,291/7,983; n_RS-II_=2,154/3,011; n_RS-III_=3,048/3,932), resulting in a final sample of 11,493 participants. Ambient air pollution (AAP) exposure was not assessed in RS, as all participants resided within a geographical area of ∼4km^2^, providing insufficient geographic contrast in exposure levels.

### Study cohorts: NELSON

Full information on the NELSON trial, including study design, study variables, nodule management protocol, and methods can be found in the original NELSON publication.^2^ Briefly, the NELSON screening trial is a population-based, randomized, controlled trial, investigating whether low-dose CT screening reduces mortality among Dutch and Belgian long-term smokers. Participants in the current study were Dutch individuals from the CT screening arm of NELSON, aged 50 to 74 and were current or former smokers (former smokers were defined as those who quit ≤10 years ago, who smoked >15 cigarettes a day for >25 years or >10 cigarettes a day for >30 years). Participants were ineligible for the current study, if they had an invalid residential ZIP code (n=35), an unknown age (n=3), or unavailable smoking data (n=25). After applying exclusion criteria, 7,364 individuals were retained for analysis.

The NELSON screening cohort was stratified into three subcohorts according to genotype data availability: ‘NELSON Ex’ (n_Ex_=2,160) lacked genotype data, ‘NELSON GenEx-I’ (n_GenEx-I_=2,896) was previously genotyped with published findings,^30^ and ‘NELSON GenEx-II’ (n_GenEx-II_=2,308) was newly genotyped for this study in 2024. Long-term exposure to AAP was assigned to the centroid of NELSON participants’ residential ZIP codes (six alphanumeric digits) via two data sources. Exposures to nitrogen dioxide (NO_2_), ozone (O_3_), and particulate matter with aerodynamic diameter of less than 10 and 2.5 micrometers (PM_10_ and PM_2.5_) were assigned based on Land Use Regression (LUR) models developed by the EXPANSE project.^31,32^ Exposure to Ultra-Fine Particles (UFP) was assigned based on Netherlands-wide LUR models which combined regional background measurements with short-term and mobile measurements.^33^

See **Supplementary Methods** for additional details on cohort description and genetic as well as AAP exposure data.

### PRS derivation, quality control, and calculation

To derive PRSs, we searched the genome-wide association studies (GWAS) Catalog for lung cancer GWAS with European populations and that included more than 10,000 lung cancer cases (to identify studies with the highest power). Large-scale multi-trait analyses were excluded. NELSON and RS were not included in the selected GWAS discovery studies.

We next applied a *P* value threshold of 1*10^-6^ to the European ancestry GWAS summary statistics and pruned the remaining variants at r^2^=0.1 using the LDlinkR R package.^36^ We kept PRSs that consisted of 10 or more independent genome-wide significant or suggestive lead variants, to have sufficient variants to calculate Gaussian PRS risk distributions. These variants were extracted from the RS and NELSON imputed datasets. Effect alleles and reported allele frequencies were harmonized and matched between the published GWAS results and the NELSON and RS genotyped datasets. PRSs from studies showing inconsistent allele frequencies with NELSON and RS and thereby hindering proper allele matching, were left out. Variants with imputation quality r^2^<0.7 in any of the three RS subcohorts, or in NELSON GenEx-I were removed from the PRS in all subcohorts.

Following the exclusion of GWAS studies and variant filtering, two PRSs remained for downstream analyses: PRS-McKay (McKay *et al.*,^35^ 48 genetic variants) and PRS-Byun (Byun *et al.*,^36^ 36 genetic variants). These variants were used consistently across all subcohorts, except in NELSON GenEx-II, where one variant in each PRS was absent in the imputed dataset (PRS-McKay: rs62621207, PRS-Byun: rs7902587). This resulted in PRS-McKay being calculated with 47 variants and PRS-Byun with 35 variants. Detailed information on the selected PRS variants is provided in **Supplementary Table S1**.

PRS-McKay and PRS-Byun were also combined into a single PRS: PRS-combined (63 genetic variants). Overlapping variants with opposing effect directions between the two studies were removed (n=5) and the effect sizes of the remaining variants were averaged (n=11). All calculated PRSs were z-transformed within RS and NELSON subcohorts.

### Statistics

The distribution of the PRSs and AAP exposures between cases and controls in RS and NELSON can be found in **Supplementary Figure S1**. The z-transformed, continuous PRS and AAP exposures were analyzed separately in univariable models. Due to the right-skewed distribution of exposure to UFP, we performed a log-transformation prior to z-transformation.

Analyses were based on available data, without imputation for missing values. Pearson correlation tests were conducted to evaluate linear relationships within the PRSs and within the AAP constituents. To calculate odds ratios (ORs) and accessory confidence intervals (CIs) we applied a generalized linear model with binomial outcome (lung cancer yes/no). Association analyses were conducted in the overall cohort and stratified by sex, age at diagnosis (<75/≥75 years), smoking status (never/former/current), histological subtype (adenocarcinoma/squamous cell carcinoma/small cell carcinoma/non-small cell carcinoma not otherwise specified), and by screening results (indeterminate and/or positive CT nodule in one of the four screening rounds). Areas under the curve (AUCs) were determined using the ‘pROC’ package.^37^ To analyze the ends of the PRS and AAP distribution, cases/controls in the bottom and top deciles were compared to cases/controls within the 40-60^th^ percentile, resembling the mean risk in each cohort. Interaction analyses were performed by applying a generalized linear model with binomial outcome, with PRS-Byun, PM_2.5_, and cigarette smoking pack-years as continuous variables, together with their multiplicative interaction terms. For all PRS analyses, fixed effects meta-analysis was performed on RS-I, RS-II, and RS-III, and NELSON GenEx-I, and NELSON GenEx-II (rmeta package). R statistical software was used to perform analyses (version 4.3.1).

## RESULTS

### Cohort characteristics

The demographic and clinical characteristics of RS and NELSON are shown in **Table 1**. Characteristics for RS and NELSON subcohorts and after stratification by case-control status can be found in **Supplementary Table S2-S3**.

**Table 1.** Demographic and clinical characteristics of RS and NELSON.

|  | **Rotterdam Study\***<br>(RS; N=11,493) | **NELSON†**<br>(N=7,364) |
| --- | --- | --- |
| **Demographics** |  |  |
| Female sex, n (%) | 6,670 (58.0) | 1,196 (16.2) |
| Age at cohort entry, years, median (IQR) | 62 (58-71) | 58 (54-62) |
| **Smoking Status at Cohort Entry, n (%)** |  |  |
| - Current | 2,382 (20.7) | 4,097 (55.6) |
| - Former | 4,914 (42.8) | 3,267 (44.4) |
| - Never | 3,882 (33.8) | .. |
| - Missing | 315 (2.7) | 0 (0.0) |
| Pack-years current smokers, median (IQR) | 30.0 (17.3-43.8) | 38.0 (28.0-49.5) |
| - Missing, n (% of current smokers) | 128 (5.4) | 0 (0.0) |
| Pack-years former smokers, median (IQR) | 15.5 (5.3-32.0) | 38.0 (29.7-49.5) |
| - Missing, n (% of former smokers) | 415 (8.4) | 0 (0.0) |
| **Environmental Exposure, mean ± SD** |  |  |
| - NO<sub>2</sub>, µg/m<sup>3</sup> | .. | 27.1 ± 6.5 |
| - O<sub>3</sub>, µg/m<sup>3</sup> | .. | 56.1 ± 4.4 |
| - PM<sub>10</sub>, µg/m<sup>3</sup> | .. | 29.2 ± 2.8 |
| - PM<sub>2.5</sub>, µg/m<sup>3</sup> | .. | 16.3 ± 1.4 |
| - UFP, particles/m<sup>3</sup> | .. | 11,477 ± 3,628 |
| **Lung Cancer Diagnoses** |  |  |
| Lung cancer cases, n (%) | 356 (3.1) | 469 (6.4) |
| Age-at-diagnosis, years, median (IQR) | 76 (69-80) | 70 (65-74) |
| Histological subtype, n (% of lung cancer cases) |  |  |
| - NSCLC | 242 (68.0) | 363 (77.4) |
| - ADN | 65 (18.3) | 244 (52.0) |
| - SQM | 84 (23.6) | 91 (19.4) |
| - NSCLC-NOS | 93 (26.1) | 28 (6.0) |
| - SCLC | 47 (13.2) | 62 (13.2) |
| - Other/NOS/unknown | 67 (18.8) | 44 (9.4) |
| **Screening Outcomes** |  |  |
| Positive/indeterminate screening result, n (%) | .. | 2,214 (30.1) |
| - Missing, n (%) | .. | 263 (3.6) |
| Lung cancer mortality, n (%) | .. | 226 (3.1) |
| All-cause mortality, n (%) | .. | 1,214 (16.5) |

Percentages are calculated as % of total unless specified otherwise. IQR: interquartile range, SD: standard deviation, NO<sub>2</sub>: nitrogen dioxide, O<sub>3</sub>: ozone, PM<sub>10</sub>: particulate matter <10 µm in diameter, PM<sub>2.5</sub>: particulate matter <2.5 µm in diameter, UFP: ultra-fine particles; <0.1 µm in diameter, ADN: adenocarcinoma, SQM: squamous cell carcinoma, SCLC: small cell lung cancer, NSCLC: non-small cell lung cancer, NOS: not otherwise specified.
\* Data represent the genotyped subset of the full RS cohort.
† **Supplementary Table S2** includes genotyped subsets of NELSON: GenEx-I and GenEx-II.

The RS cohort (N=11,493) consisted predominantly of female participants (n=6,670; 58.0%), with a median age at baseline of 63 years. Participants were often former smokers (n=4,914; 42.8%,), and the median number of pack-years of former smokers was 15.5, compared to 30.0 for current smokers. During 29, 18, and 12 years of follow-up (RS-I, RS-II, and RS-III, respectively), 356 participants developed lung cancer (3.1%). Non-small cell lung cancer (NSCLC) was the most common histological subtype (n=242; 68.0% of cases). Due to historical diagnostic practices, many lung cancer cases from the earlier recruitment rounds of the RS (initiated in 1989) were classified as NSCLC not otherwise specified (NSCLC-NOS) or lacked a recorded histological subtype (n=160; 44.9%). The genotyped RS participant characteristics did not differ from the full cohort (**Supplementary Table S2**).

In contrast to RS, NELSON (N=7,364) was mostly male (n=6,168; 83.8%), especially NELSON GenEx-I (n=2,864; 98.9%; **Supplementary Table S2**). Median age at randomization was 58 years. Compared to RS, NELSON participants were more often current smokers (n_NELSON_=4,097; 55.6% vs. n_RS_=2,382; 20.7%) with a higher median pack-year history (38.0 years for former and current smokers), reflecting the NELSON inclusion criteria. AAP exposures ranged from 10.2 to 53.5 µg/m^3^ for NO_2_, 33.1 to 75.2 µg/m^3^ for O_3_, 22.0 to 40.4 µg/m^3^ for PM_10_, 12.7 to 20.3 µg/m^3^ for PM_2.5_, and 7,435 to 76,723 particles/m^3^ for UFP. We found strong, inverse correlations between O_3_ and all other AAP constituents (-0.79<r<-0.66), and moderate to strong positive correlations between NO_2_, PM_10_, PM_2.5_, and UFP (0.43<r<0.79) (**Supplementary Figure S2**).

In total, 2,214 NELSON participants (30.1%) received an indeterminate or positive screening result in at least one of the four screening rounds. During 13 years of follow-up, 469 participants developed lung cancer (6.4%), of which most participants developed NSCLC (n=363; 77.4% of cases). Lung cancer incidence was higher in the NELSON Ex subcohort (n=188; 8.7%; **Supplementary Table S2**). Median age-at-diagnosis was lower in NELSON compared to RS (70 vs. 76), reflecting the younger age at cohort entry, the elevated baseline risk in this population, and the targeted nature of the screening trial, facilitating early detection. During follow-up, 1,214 (16.5%) of the 7,364 NELSON participants died from all causes and 226 (3.1%) participants died from lung cancer.

### Associations with lung cancer and univariable discriminative ability

We derived two European-oriented PRSs from GWAS variants: PRS-McKay and PRS-Byun (48 and 36 independent variants, respectively; see Methods). PRS-Byun showed a positive association with lung cancer and modest discriminative ability in both RS and NELSON (OR_RS_ [95%-CI]: 1.19 [1.07-1.32], AUC_RS_ [95%-CI]: 0.541 [0.510-0.572]; OR_NELSON_ [95%-CI]: 1.28 [1.13-1.44], AUC_NELSON_ [95%-CI]: 0.569 [0.533-0.605]; **Figure 2**). Similar results were obtained with PRS-McKay, although associations with lung cancer were weaker, particularly in RS (OR_RS_ [95%-CI]: 1.09 [0.98-1.21], AUC_RS_ [95%-CI]: 0.523 [0.492-0.554]; OR_NELSON_ [95%-CI]: 1.22 [1.08-1.37], AUC_NELSON_ [95%-CI]: 0.560 [0.524-0.595]). PRS analyses stratified by sex, age-at-diagnosis, smoking status, and other available histological subtypes showed consistent positive associations for PRS-Byun across RS and NELSON (**Supplementary Figure S3**). PRS-McKay also showed positive associations in most strata. The results for separate PRS analyses of RS-I, RS-II, and RS-III, NELSON GenEx-I, and GenEx-II are presented in **Supplementary Table S4**. Sensitivity analyses restricted to incident cases and histologically confirmed cases in RS yielded results consistent with the primary analysis including all cases (**Supplementary Table S5-S6**).

**Figure 2.**
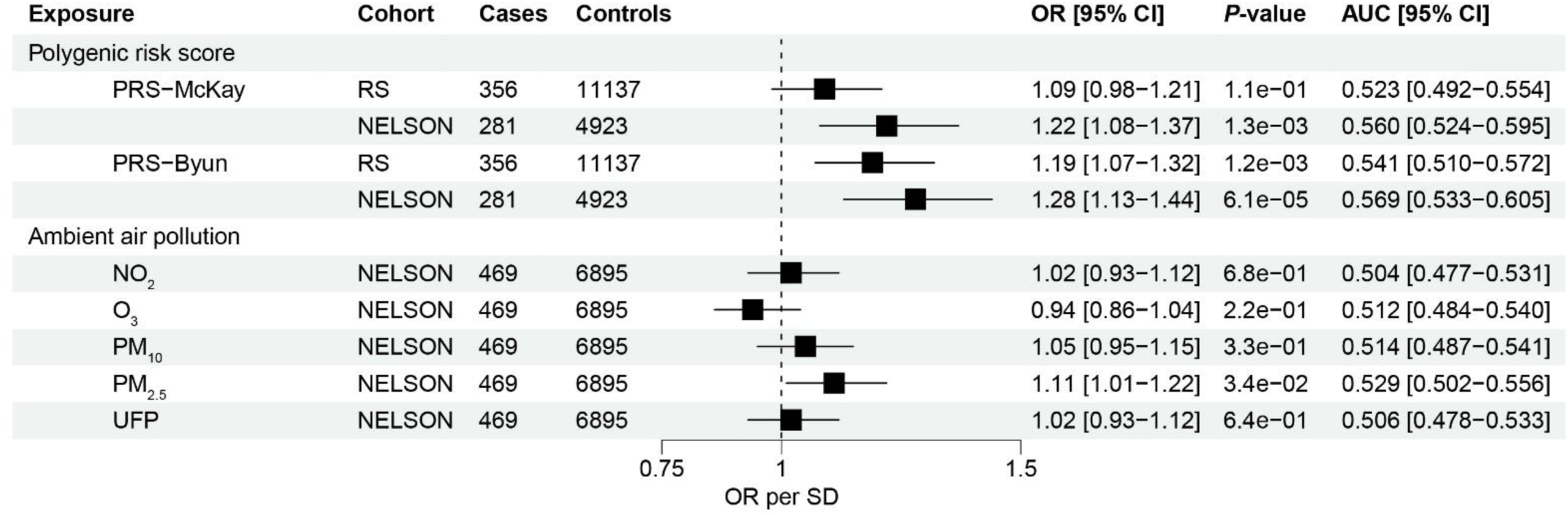
Effect estimates and univariable discrimination of PRSs and AAP constituents. Generalized linear univariable model analyzing the PRSs and AAP exposures, with binomial outcome: lung cancer yes/no. The PRS analyses represent a fixed effects meta-analysis on RS-I, RS-II, and RS-III, and on NELSON GenEx-I, and GenEx-II. The OR corresponds to the increase in odds of lung cancer per standard deviation increase in the PRS and AAP exposures. LC: lung cancer, OR: odds ratio, SD: standard deviation, AUC: area under the curve, NO_2_: nitrogen dioxide, O_3_: ozone, PM_10_: particulate matter <10 μm in diameter, PM_2.5_: particulate matter <2.5 μm in diameter, UFP: ultra-fine particles; <0.1 μm in diameter.

Between PRS-McKay and PRS-Byun, 16 variants overlapped directly or were in moderate linkage disequilibrium (r^2^>0.3). The Pearson correlation between the two PRSs ranged from 0.62 to 0.64 in RS and NELSON subcohorts, respectively. Analyzing both PRSs in one model yielded slightly diminished effect sizes for PRS-McKay (OR_RS_ [95%-CI]: 0.96 [0.84-1.10], OR_NELSON_ [95%-CI]: 1.07 [0.92-1.25]), while the effect sizes for PRS-Byun remained stable (OR_RS_ [95%-CI]: 1.22 [1.07-1.40], OR_NELSON_ [95%-CI]: 1.22 [1.05-1.43]). Hence, a combined PRS did not yield a stronger association between the PRS and lung cancer (**Supplementary Table S7**).

PM_2.5_ was associated with all-type lung cancer in NELSON (OR per SD [95%-CI]: 1.11 [1.01-1.22], AUC [95%-CI]: 0.529 [0.502-0.556]) (**Figure 2**). In histological subtype analysis, positive associations were observed for lung adenocarcinoma with NO_2_, PM_10_, PM_2.5_ and UFP (OR_NO2_ [95%-CI]: 1.13 [1.00-1.29], OR_PM10_ [95%-CI]: 1.16 [1.01-1.32], OR_PM2.5_ [95%-CI]: 1.14 [1.00-1.29], OR_UFP_ [95%-CI]: 1.16 [1.04-1.30]; **Supplementary Figure S4**). Positive associations were also observed for NSCLC-NOS, similar to our findings for adenocarcinoma (**Supplementary Figure S4**). No positive association was found for squamous and small cell lung carcinoma. NO_2_, PM_10_, PM_2.5_, and UFP showed a trend toward a positive association only in men (**Supplementary Figure S4**).

To explore the potential influence of AAP exposure timing on lung cancer risk, we conducted a secondary analysis using estimated pollutant exposures three years prior to randomization. No associations were found between earlier exposure estimates and lung cancer (**Supplementary Table S8**).

### Top and bottom decile analysis

To assess whether risk-increasing effects scaled with levels of PRS or AAP exposure, individuals within the highest and lowest deciles of the PRSs and AAP distributions were compared with a reference group representing average exposure (i.e., the 40^th^-60^th^ percentiles) within the study population. This revealed an asymmetric relationship between the PRSs and lung cancer diagnosis (**Supplementary Figure S5**). The top deciles of the PRSs generally showed positive effects, particularly the top decile of PRS-Byun in NELSON (OR [95%-CI]: 2.40 [1.57-3.67], AUC [95%-CI] 0.605 [0.543-0.668]), while effect sizes at the bottom deciles remained close to 1.

Similarly, for the AAP exposures, an asymmetric relationship for PM_2.5_ and NO_2_ deciles was observed, with a stronger risk-increasing effect in the top decile compared to modest or null effects in the bottom decile (**Supplementary Figure S5**). For O_3_, the bottom decile showed a positive association with lung cancer (OR [95%-CI]: 1.47 [1.03-2.08], AUC [95%-CI]: 0.545 [0.502-0.587]). Overall, we did not observe uniform trends for PM_10_ and UFP.

### Discriminative ability on top of pack-years, age, and sex

Next, we analyzed the PRSs and AAP constituents in a multivariable model in NELSON (**Table 2**). Due to the correlations between PRS-McKay and PRS-Byun, and between AAP constituents, only one PRS and one AAP constituent was included in the model. PRS-Byun and PM_2.5_ were selected based on their strongest univariable association with lung cancer when treated as continuous variables. A baseline model including age, sex, and pack-year history was constructed, to which PRS-Byun and PM_2.5_ were introduced to evaluate their impact on the model’s discriminative ability.

**Table 2.** Discriminative ability of models incorporating PRS and PM_2.5_.

| Model* | Variable | OR<sub>variable</sub> [95% CI] | AUC<sub>model</sub> [95% CI] |
| --- | --- | --- | --- |
| **Univariable model** |  |  |  |
| PRS-Byun | PRS-Byun | 1.28 [1.13-1.44] | 0.569 [0.533-0.605] |
| PM<sub>2.5</sub> | PM<sub>2.5</sub> | 1.08 [0.97-1.21] | 0.527 [0.492-0.562] |
| **Baseline model** |  |  |  |
| Age + sex + pack-years | - | - | 0.643 [0.608-0.679] |
| **Baseline model and PRS or AAP** |  |  |  |
| Baseline + PRS-Byun | PRS-Byun | 1.27 [1.13-1.44] | 0.655 [0.620-0.691] |
| Baseline + PM<sub>2.5</sub> | PM<sub>2.5</sub> | 1.08 [0.96-1.21] | 0.646 [0.610-0.681] |
| **Full model** |  |  |  |
| Baseline + PRS-Byun + PM<sub>2.5</sub> | PRS-Byun | 1.27 [1.13-1.44] | 0.659 [0.624-0.695] |
| " | PM<sub>2.5</sub> | 1.07 [0.96-1.21] | " |

Variables were analyzed in a generalized linear model with binomial outcome: lung cancer yes/no. The number of pack-years smoked was z-transformed and added to the model as a continuous variable. The analyses represent a fixed effects meta-analysis on NELSON GenEx-I and GenEx-II. The OR corresponds to the increase in odds of lung cancer per standard deviation increase in the PRS and AAP exposures. OR: odds ratio, AUC: area under the curve, LC: lung cancer, PM<sub>2.5</sub>: particulate matter <2.5 µm in diameter.
\* The number of cases and controls was identical across all analyses: n<sub>cases</sub>=281, n<sub>controls</sub>=4,923.

To allow for analysis of PRS-Byun and PM_2.5_ within the same model, we meta-analyzed PM_2.5_ in the NELSON GenEx-I and GenEx-II subcohorts. This reduced the sample size of the observed PM_2.5_ association (n=281 vs. n=469 lung cancer cases), slightly diminishing its discrimination in this subset (AUC [95%-CI] 0.527 [0.492-0.562] vs. 0.529 [0.502-0.556], respectively; **Table 2** and **Figure 2**).

A baseline model with only age, sex and pack-years yielded an AUC of 0.643 (95%-CI: 0.608-0.679; **Table 2**). The addition of PRS-Byun and PM_2.5_ increased the AUC by 0.012 and 0.003, respectively. Incorporating both PRS-Byun and PM_2.5_ further improved the model’s discriminatory performance, yielding an AUC of 0.659 (95%-CI: 0.624-0.695). The associations of PRS-Byun and PM2.5 with lung cancer remained consistent after adjustment for age, sex, and pack-years (OR_PRS-Byun_ [95%-CI]: 1.27 [1.13-1.44] vs. 1.28 [1.13-1.44], OR_PM2.5_ [95%-CI]: 1.07 [0.96-1.21] vs. 1.08 [0.97-1.21]; **Table 2**). The multiplicative interaction terms between PRS-Byun and pack-years, PM_2.5_ and pack-years, and PRS-Byun and PM_2.5_ were not statistically significant (*P*-value: 0.77, 0.12, and 0.63, respectively).

### Associations with screening outcomes

In order to assess usefulness for risk stratification in the screening process, we analyzed the relationship between exposures and lung cancer in individuals with at least one indeterminate or positive CT result, requiring additional follow-up such as short-term repeat CT scans or pulmonologist referral for further (diagnostic) workup. We also examined the relationship between exposures and key screening outcomes, including cause-specific mortality (lung cancer and non-lung cancer) and indeterminate or positive screening CT results, to determine whether the PRSs and AAP capture competing mortality risks or general susceptibility to pulmonary nodules, respectively.

We noted a positive association between PRS-McKay and a positive/indeterminate screening result (OR [95%-CI]: 1.08 [1.02-1.15]; **Figure 3**). This association persisted when we restricted the analysis to NELSON participants who did not develop lung cancer during study follow-up (OR [95%-CI]: 1.07 [1.01-1.14]; **Supplementary Table S9**). Restricting analysis to participants with at least one indeterminate or positive CT result yielded a consistent positive association with lung cancer for PRS-Byun (OR [95%-CI]: 1.27 [1.09-1.48]), while PRS-McKay showed a slightly attenuated association (OR [95%-CI]: 1.13 [0.97-1.32]). Overall, effect sizes in this subgroup were similar in magnitude and direction to those in the full NELSON cohort, but with broader confidence intervals due to the smaller sample size. PRS-McKay and PRS-Byun were associated with lung cancer mortality (OR [95%-CI]: 1.24 [1.05-1.47] for both), but not with non-lung cancer mortality. The AAP constituents were not associated with mortality.

**Figure 3.**
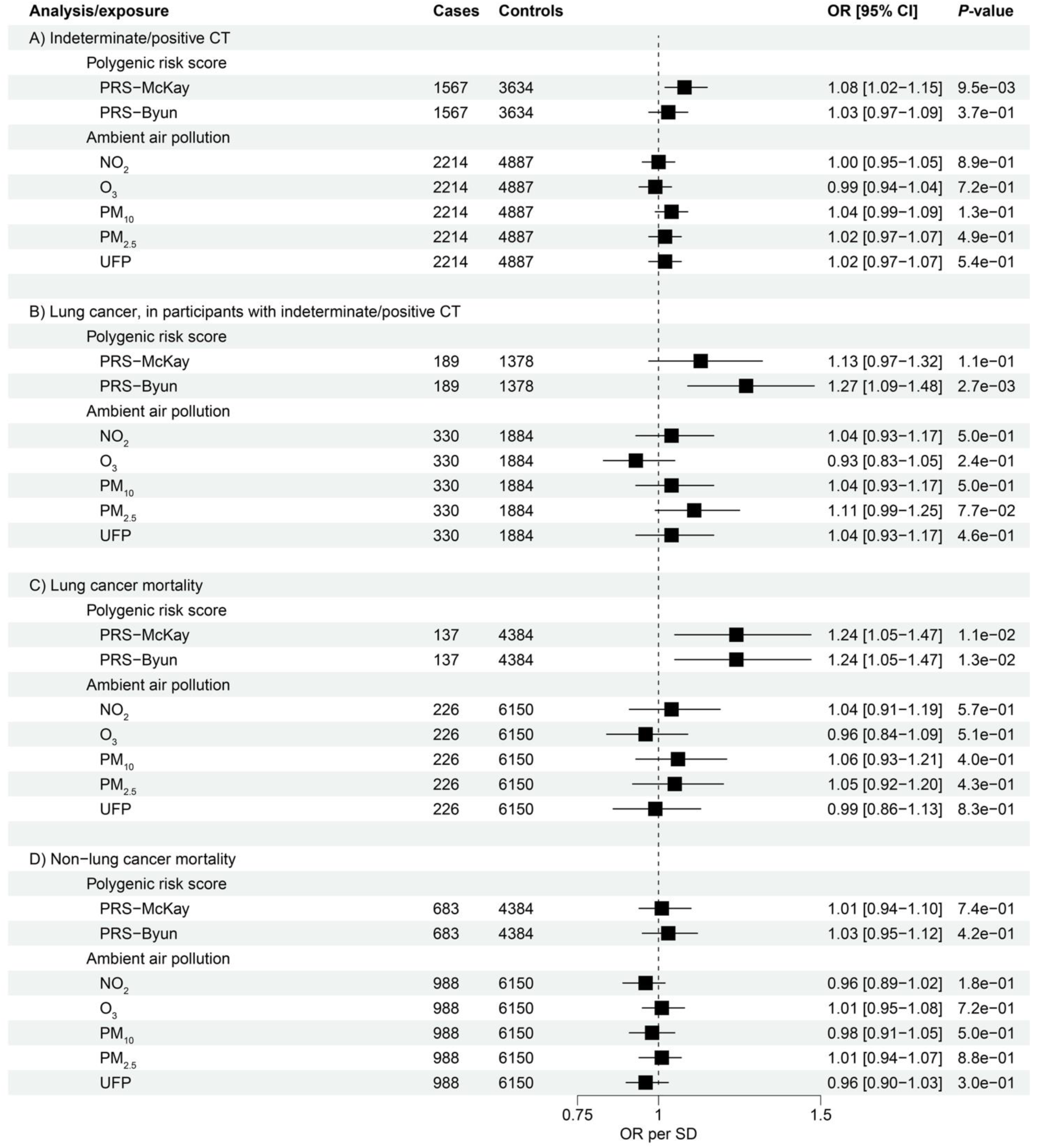
Associations of PRSs and AAP constituents with screening outcomes. Generalized linear univariable model analyzing the PRSs and AAP constituents, with binomial outcomes: A) indeterminate/positive CT result in one of the four screening rounds, requiring follow-up imaging or clinical referral, yes/no; B) lung cancer yes/no, stratified by an indeterminate/positive CT result in one of the four screening rounds; C) and D) all-cause mortality yes/no stratified by C) lung cancer-specific mortality and D) non-lung cancer mortality. The PRS analyses represent a fixed effects meta-analysis on NELSON GenEx-I, and GenEx-II. The OR corresponds to the increase in odds of the outcome per standard deviation increase in the PRS and AAP exposures. LC: lung cancer, OR: odds ratio, SD: standard deviation, NO_2_: nitrogen dioxide, O_3_: ozone, PM_10_: particulate matter <10 μm in diameter, PM_2.5_: particulate matter <2.5 μm in diameter, UFP: ultra-fine particles; <0.1 μm in diameter.

## DISCUSSION

In the current paper, we find that in the NELSON lung cancer screening cohort of high-risk individuals, our two PRSs (PRS-McKay and PRS-Byun), and ambient PM_2.5_ at residential ZIP code level were associated with lung cancer risk. PRS-Byun modestly added discriminative ability beyond age, sex, and smoking history, with participants in the top decile of PRS-Byun exhibiting a 2.4-fold increased risk compared to those in the mid-distribution range. Subtype analyses further revealed that ambient PM and particularly UFP were associated with lung adenocarcinoma, the subtype with the highest global incidence.^28^ Collectively, these findings warrant further investigation for the potential clinical relevance of PRS and exposure to PM in lung cancer screening.

Our results extend prior evidence showing an association between PRS with and of lung cancer in general and high-risk populations.^8–12,21^ Our per-SD OR estimates of 1.1-1.3 are broadly consistent with previously reported lung cancer PRS per-SD effect sizes, which fall between 1.2 and 1.4.^10,12^ For both PRS-McKay and PRS-Byun, we observed asymmetric associations with odds of lung cancer: a higher relative score was associated with increased odds, whereas a lower score did not associate with lower odds. Individuals in the top decile of the PRS distribution had ORs between 1.3 and 2.4 when compared with those representing average genetic risk (40^th^-60^th^ percentile). Prior studies typically compare more extreme PRS categories, such as the top decile versus the bottom decile, and report effect estimates in the 1.5-3.5 range.^8–10,12^ A study by McHugh *et al.* found that invitation for prostate cancer screening based on a top decile PRS, identified 103 clinically significant cancers in 468 screened men, 72% of which would not have met criteria for biopsy under the current UK PSA/MRI criteria.^38^ PRS-Byun exhibits similar univariable performance in the top decile as the prostate cancer PRS used in McHugh *et al.* (i.e. OR_PRS-Byun_: 2.4, AUC_PRS-Byun_: 0.61 vs. OR_PRS-McHugh_: 2.0, AUC_PRS-McHugh_: 0.55).^38^ These findings (including those presented here) collectively suggest that applying a cut-off value on the higher range of the PRS distribution may help refine selection criteria for lung cancer screening.

In a multivariable model, PRS-Byun demonstrated an AUC improvement of 0.012 beyond age, sex, and pack-years in the NELSON cohort. These findings are consistent with the high-risk case-control analysis by Lebrett *et al.*, in which incorporating various published lung cancer PRSs into the PLCOm2012-based screening model increased the AUC by between 0.002 and 0.015.^11^ Whether such modest improvements in AUC translate into clinically meaningful or cost-effective changes in lung cancer screening practice remains uncertain and will require further study.

To move beyond global discrimination metrics, we assessed the associations of the PRS and PM_2.5_ with screening outcomes, as markers of their potential clinical relevance. We observed that PRS-McKay was associated with indeterminate or positive CT results irrespective of a subsequent lung cancer diagnosis (OR: 1.1), whereas PRS-Byun was not. These differences suggest that PRS-McKay also captures odds of pulmonary nodule formation detectable on CT, possibly reflecting partially distinct underlying biology or GWAS phenotype definitions. Notably, PRS-McKay and PRS-Byun were both associated with lung cancer-specific mortality (OR: 1.2), but not with other prevalent causes of death in this population, such as cardiovascular disease. This result mirrors findings from the National Lung Screening Trial, where a lung cancer PRS was independently associated with lung cancer incidence and mortality, but did not relate to non-lung cancer or all-cause mortality.^21^ This reinforces that, unlike smoking-based criteria, PRS has the potential to refine selection among screening-eligible individuals toward those with higher odds of dying from lung cancer specifically.

Multiple large cohort and meta-analytic studies have found an association between PM_2.5_ exposure and lung cancer. Meta-analyses of prospective cohorts report relative risks around 1.1-1.2 per 10 µg/m^3^ increase in PM_2.5_ for lung cancer incidence or mortality, with the European Study of Cohorts for Air Pollution Effects (ESCAPE) study reporting an effect size of about 1.2 per 5 µg/m^3^ for overall lung cancer and 1.5 per 5 µg/m^3^ for adenocarcinoma.^13,14,17^ Our estimated OR of 1.1 per SD increase in PM_2.5_ is broadly consistent with these findings. We also found slightly higher estimates for PM_2.5_, and other AAP constituents in relation to lung adenocarcinoma, consistent with the pattern of stronger associations for this histologic subtype reported in the ESCAPE study and by Hamra *et al*.^13,14^ Of note, our results suggest that exposure to PM constituents, even when estimated at baseline at the ZIP code level, is associated with odds of lung cancer and adenocarcinoma, supporting its feasibility for use in risk stratification models.

Notably, we identified a link between UFP exposure and increased odds of adenocarcinoma. In contrast to PM_2.5_ and PM_10_, UFP levels remain largely unregulated and the impact of UFP exposure on health outcomes remains relatively understudied. This is mainly due to limited long-term exposure data, high spatial and temporal variability, and the absence of standardized monitoring protocols. Consequently, few prospective studies have investigated UFPs in relation to cancer risk.^27,39,40^ Our findings contribute novel evidence from a high-risk screening population linking UFP to lung cancer and warrant further large-scale, multi-pollutant studies to confirm and contextualize these associations.

We observed an unexpected inverse association between ambient O_3_ and lung cancer incidence in a screening population. In our data, O_3_ was strongly and inversely correlated with all other AAP constituents, suggesting that the apparent protective association may reflect lower concentrations of other pollutants in areas with higher O_3_. Supporting this notion, other studies reported an inverse association between O_3_ and lung cancer mortality,^41,42^ which was attenuated after adjustment for NO_2_.^41^

Limitations of our study include the following. First, we currently lack data on several known risk factors, including BMI, family history of lung cancer, occupational hazards, COPD diagnosis, education, and socio-economic status, limiting our ability to construct a more comprehensive risk model in NELSON. However, previous studies have shown that PRSs and PM exposure associate with lung cancer risk largely independent of these variables.^9–14,17,20^ Second, our cohort consisted predominantly of individuals of European ancestry. Although lung cancer PRSs have also shown potential for risk stratification in Chinese populations^8^, further validation in diverse cohorts is needed—requiring GWAS studies to also include such ancestries. Third, the limited number of women in NELSON restrict the generalizability of our findings for AAP. The observed trend toward a positive association between AAP constituents and lung cancer only in men, alongside evidence suggesting sex-specific differences in AAP-related lung cancer risk,^43^ warrants further investigation. Fourth, our PRSs were derived from GWAS studies in populations that may differ slightly from the Rotterdam Study and/or NELSON cohorts. Future lung cancer GWAS studies in larger and more tailored populations might offer improved PRS performance. Additionally, we utilized a relatively straightforward pruning & thresholding PRS construction, to evaluate the value of PRS in concept, and more sophisticated approaches exist to optimize performance in specific applications. Finally, exposure misclassification is possible for the AAP estimates due to the reliance on baseline ZIP codes, and the absence of data on residential mobility of NELSON participants during the thirteen years of follow-up. Exposure misclassification will particularly affect UFP exposure estimates as they were assigned several years after baseline. Indeed, in a similar study, sensitivity analyses restricted to non-movers reported stronger associations between PM_10_ and PM_2.5_ and lung cancer,^13^ suggesting that we may underestimate the true effect of PM in this study.

In conclusion, this study found that lung cancer PRS and exposure to PM were each associated with lung cancer in a high-risk screening cohort, with both evaluated PRSs additionally associated with lung cancer mortality. These findings suggest potential relevance of genetic and environmental risk factors for risk-stratified screening. Although discriminative ability was only modestly improved upon adding these exposures to a baseline model, future studies should evaluate the association of PRS with lung cancer-specific mortality and its utility in identifying individuals most likely to benefit from screening.

## DATA AVAILABILITY

The R code used for data processing and analysis in this study is available from the corresponding authors upon reasonable request. Researchers interested in using The Rotterdam Study data must submit a formal data access request to the cohort’s management team via email. Access to the NELSON study data is controlled by the NELSON study Data Access Board (see https://umcgresearchdatacatalogue.nl/all/cohorts/NELSON).

## ACKNOWLEDGMENTS

The authors would like to thank the NELSON-POP consortium team, the Rotterdam Study team, and staff of the Department of Pulmonary Medicine at the Erasmus MC University Medical Center for their invaluable input and support.

## AUTHOR CONTRIBUTIONS

**Lianne Trap:** Conceptualization; Data curation; Formal analysis; Investigation; Roles/Writing - original draft; Writing - review & editing

**Ramazan Büyükcelik:** Investigation

**Noa Antonissen:** Data curation; Writing - review & editing

**Grigory A. Sidorenkov:** Data curation; Writing - review & editing

**Rikje Ruiter:** Data curation; Resources; Writing - review & editing

**Jolande van Heemst:** Data curation; Resources

**Bahar Sedaghati-Khayat:** Methodology; Writing - review & editing

**Bernard S. Stikker:** Methodology; Writing - review & editing

**Daphne W. Dumoulin:** Data curation; Writing - review & editing

**Hester A. Gietema:** Writing - review & editing

**Marjolein A. Heuvelmans:** Writing - review & editing

**Firdaus A.A. Mohamed Hoesein:** Writing - review & editing

**Pim A. de Jong:** Writing - review & editing

**André G. Uitterlinden:** Writing - review & editing

**Guy Brusselle:** Writing - review & editing

**Colin Jacobs:** Writing - review & editing

**Joachim G.J.V. Aerts:** Writing - review & editing

**Roel C.H. Vermeulen:** Resources; Writing - review & editing

**Geertruida H. De Bock:** Writing - review & editing

**Harry J.M. Groen:** Writing - review & editing

**Rozemarijn Vliegenthart:** Funding acquisition; Project administration; Writing - review & editing

**George S. Downward:** Conceptualization; Formal analysis; Investigation; Methodology; Supervision; Roles/Writing - original draft; Writing - review & editing

**Ralph Stadhouders:** Funding acquisition; Conceptualization; Investigation; Project administration; Supervision; Roles/Writing - original draft; Writing - review & editing

**Jeroen van Rooij:** Conceptualization; Formal analysis; Investigation; Methodology; Supervision; Roles/Writing - original draft; Writing - review & editing

## ETHICS DECLARATION

The Rotterdam Study has been approved by the Medical Ethics Committee of the Erasmus MC (registration number MEC 02.1015) and by the Dutch Ministry of Health, Welfare and Sport (Population Screening Act WBO, license number 1071272-159521-PG). The Rotterdam Study Personal Registration Data collection is filed with the Erasmus MC Data Protection Officer under registration number EMC1712001. The Rotterdam Study has been entered into the Netherlands National Trial Register (www.trialregister.nl) and into the World Health Organization International Clinical Trials Registry Platform (www.who.int/ictrp/network/primary/en/) under shared catalogue number NTR6831. All participants provided written informed consent to participate in the study and to have their information obtained from treating physicians.

The NELSON trial was approved by the Dutch Minister of Health and by the Medical Ethics Committee. All participants in the study provided informed consent for participation in the trial and for linkage to their data.

## DECLARATION OF INTERESTS

R.V. is supported by institutional research grants from Siemens Healthineers, and has received honorarium for lectures from Bayer Healthcare, Siemens Healthineers, and Wiley. D.W.D. has received personal and institutional consulting fees from MSD, Amgen, Roche, BMS, AstraZeneca and Pfizer, all unrelated to the submitted work. The Department of Radiology at UMC Utrecht receives research support from Philips Healthcare, unrelated to the submitted work. All other authors declare no competing interests.

### NELSON-POP consortium members

Joachim G. Aerts, MD, PhD, University Medical Center Rotterdam, Erasmus University Rotterdam, Rotterdam, The Netherlands

Robin Cornelissen, MD, PhD, University Medical Center Rotterdam, Erasmus University Rotterdam, Rotterdam, The Netherlands

Ralph Stadhouders, PhD, University Medical Center Rotterdam, Erasmus University Rotterdam, Rotterdam, The Netherlands

Jeroen G.J. van Rooij, PhD, University Medical Center Rotterdam, Erasmus University Rotterdam, Rotterdam, The Netherlands

Lianne Trap, M.Sc., University Medical Center Rotterdam, Erasmus University Rotterdam, Rotterdam, The Netherlands

Kristiaan Nackaerts, MD, PhD, University Hospital Leuven, Catholic University of Leuven, Leuven, Belgium

Walter de Wever, MD, PhD, University Hospital Leuven, Catholic University of Leuven, Leuven, Belgium

Hester A. Gietema, MD, PhD, Maastricht University Medical Center, Maastricht University, Maastricht, The Netherlands

Mathias Prokop, MD, PhD, Radboud University Medical Center, Nijmegen, The Netherlands

Cornelia Schaefer-Prokop, MD, PhD, Radboud University Medical Center, Nijmegen, The Netherlands

Colin Jacobs, PhD, Radboud University Medical Center, Nijmegen, The Netherlands

Noa Antonissen, MD, Radboud University Medical Center, Nijmegen, The Netherlands

Geertruida H. de Bock, PhD, University Medical Center Groningen, University of Groningen, Groningen, The Netherlands

Marjolein A. Heuvelmans, MD, PhD, University Medical Center Groningen, University of Groningen, Groningen, The Netherlands

Grigory Sidorenkov, PhD, University Medical Center Groningen, University of Groningen, Groningen, The Netherlands

Danrong Zhong, M.Sc., University Medical Center Groningen, University of Groningen, Groningen, The Netherlands

Harry J.M. Groen, MD, PhD, University Medical Center Groningen, University of Groningen, Groningen, The Netherlands

Rozemarijn Vliegenthart, MD, PhD, University Medical Center Groningen, University of Groningen, Groningen, The Netherlands

Nils van der Velden, M.Sc., University Medical Center Groningen, University of Groningen, Groningen, The Netherlands

Pim A. de Jong, MD, PhD, University Medical Center Utrecht, Utrecht University, Utrecht, The Netherlands

Firdaus A. A. Mohamed Hoesein, MD, PhD, University Medical Center Utrecht, Utrecht University, Utrecht, The Netherlands

Stijn Bunk, M.Sc., University Medical Center Utrecht, Utrecht University, Utrecht, The Netherlands

George S. Downward, PhD, University Medical Center Utrecht, Utrecht University, Utrecht, The Netherlands Roel

C.H. Vermeulen, PhD, University Medical Center Utrecht, Utrecht University, Utrecht, The Netherlands

