## Supplementary Data for "Evaluation of polygenic risk scores and ambient air pollutants for lung cancer risk stratification in a lung cancer screening cohort"

### SUPPLEMENTARY DATA CONTENTS

Supplementary Methods ... 3
Study cohorts: the Rotterdam Study (RS) ... 3
Study cohorts: NELSON ... 3
Supplementary Table S1. PRS variants and imputation in RS and NELSON. ... 6
Supplementary Figure S1. Distributions of exposures between lung cancer cases and controls. 9
Supplementary Table S2. RS and NELSON subcohort demographics and clinical characteristics. ... 10
Supplementary Table S3. Subcohort characteristics split by case-control status. ... 11
Supplementary Figure S2. Pearson correlations (r) between ambient air pollution constituents in NELSON. ... 12
Supplementary Figure S3. Stratified analyses of continuous PRSs in RS and NELSON. ... 13
Supplementary Table S4. Subcohort PRS results of main association analysis. ... 14
Supplementary Table S5. Sensitivity analysis for incident cases in RS. ... 15
Supplementary Table S6. Sensitivity analysis for histologically confirmed cases in RS. ... 16
Supplementary Table S7. Combined PRS performance across RS subcohorts. ... 17
Supplementary Figure S4. Stratified analysis of ambient air pollution constituents in NELSON. 18
Supplementary Table S8. Analysis of pollutant exposures three years prior to randomization. . 19
Supplementary Figure S5. Top and bottom deciles of PRS and ambient air pollution constituents. ... 20
Supplementary Table S9. Association of PRSs and AAP constituents with indeterminate/false-positive CT results. ... 21

### Supplementary Methods

#### Study cohorts: the Rotterdam Study (RS)

*Cohort description*

The RS is a Dutch, prospective, mid-late life cohort, which primarily investigates diseases of old age. Briefly, RS consists of multiple subcohorts, each of which contains inhabitants of the Ommoord suburb in Rotterdam: RS-I, RS-II, and RS-III. RS-I and II were aged 55 and over, and recruited in 1989 and 2000. RS-III participants were over 45 years of age and recruited in 2006. Information related to lung cancer and histological subtype, was obtained up until December 13$^{th}$, 2018 via medical records, hospital discharge letters, linkage with the national hospital discharge registry (PALGA), and linkage with the Netherlands Cancer Registry (NCR), with confirmation by radiological and/or histological assessment. The primary genetic analyses included all lung cancer cases (n=356). To assess the robustness of our findings, sensitivity analyses were conducted restricting to incident cases only (n=339) as well as to histologically confirmed cases only (n=311). Smoking history was self-reported at baseline via questionnaires.

*Genetic data*

RS participants were genotyped on the Illumina 550K array (RS-I and RS-II) and the Illumina 610K array (RS-III). Variants were imputed against the 1000 Genomes reference panel and reported on genome build GRCh37. Participants of RS are primarily of European ancestry (>96%). Therefore, analyses were restricted to RS participants of genotypic European ancestry. Participants who were related were left out within RS subcohorts, but not between subcohorts.

#### Study cohorts: NELSON

*Cohort description*

The NELSON screening trial is a population-based, randomized, controlled trial, investigating whether low-dose CT screening reduces mortality among Dutch and Belgian long-term smokers. Four rounds of low-dose CT screening were conducted, at baseline and at years 1, 3, and 5.5. Nodules were assessed based on volume and volume doubling time and were labelled negative, indeterminate, or positive. Participants with an indeterminate screening result received a follow-up scan before the next screening round and participants with a positive screening result were referred to a pulmonologist for further (diagnostic) workup.

Clinical data, including lung cancer incidence, histological subtype, stage, and cause of death were obtained through linkages with the Dutch Center for Genealogic and Heraldic Studies,

Statistics Netherlands, and the Netherlands Cancer Registry (NCR). Because of registry linkage, loss to follow-up is expected to be minimal and was not further analyzed. Smoking data was self-reported, with smoking status and pack-years reported at baseline. Other risk factors for lung cancer, such as BMI, family history of lung cancer, or occupational hazards, were not available.

*Genetic data*

NELSON participants in GenEx-I and GenEx-II were genotyped on the Illumina 610K-Quad array and the Illumina GSA-MD array, respectively. Variants were imputed against the 1000 Genomes reference panel and reported on genome build GRCh37. As the NELSON cohort consists mostly of individuals with European ancestry (98% of genotypes), PRS analysis were again restricted to only participants of (genotypic) European ancestry. Samples were excluded based on the following criteria:

- Missing reported or genotyped sex (n=109)
- Mismatch between reported and genotyped sex (n=8)
- Missing variants >2.5% (n=14)
- Heterozygosity and homozygosity outliers (n=21)
- Genetic duplicates and first- or second-degree familial relationships (n=31)
- Non-European ancestry based on principle component analysis (n=50)

Variants were excluded based on the following criteria:

- Low call rate <97.5% (n=16,602)
- Low call rate after zCall <99% (n=31)
- Failed Hardy-Weinberg equilibrium test $P \leq 1*10^{-5}$ (n=509)

*Ambient air pollution (AAP) exposure*

Long-term exposure to AAP was assigned to the centroid of NELSON participants' residential ZIP codes (six alphanumeric digits) via two data sources.

Exposures to nitrogen dioxide ($NO_2$), ozone ($O_3$), and particulate matter with aerodynamic diameter of less than 10 and 2.5 micrometers ($PM_{10}$ and $PM_{2.5}$) were assigned based on Land Use Regression (LUR) models developed by the EXPANSE project. Briefly, routine monitoring stations from the European Environmental Agency were collected and aggregated to long-term annual averages (2000 to 2019). Diverse spatial predictor variables, such as roads, satellite data, and chemical transport models, were utilized for geographically weighted regression, deriving annually averaged exposure models at a 25m-by-25m grid across Europe for the years 2000 to 2019. Cross validation of these models showed an $R^2$ of 0.66 for $NO_2$, 0.58 for $O_3$, 0.62 for $PM_{10}$, and 0.77 for $PM_{2.5}$. The annual average of AAP at the year of randomization, and the average of the three years prior to randomization were extracted for each NELSON participant and assigned as markers of long-term exposure.

Exposure to Ultra-Fine Particles (UFP) was assigned based on Netherlands-wide LUR models which combined regional background measurements with short-term and mobile measurements. Using an electric car, UFP concentrations were measured across the Netherlands over a 14-month period (2016 to 2017). Measurements were collected three times and concentrations were averaged per road segment. These mobile measurements were supplemented by regional background monitoring data (collected from 20 sites with three measurement periods each lasting two weeks at a time), which was used to derive annual average background concentrations. LUR modelling was performed via supervised stepwise regression, also undergoing a 'deconvolution' method, where the average concentration at each road segment was segregated into a local and background signal. Long-term model validation showed an $R^2$ of 0.60 for this approach. Exposures from this model were assigned as a 'best available' estimate.

### Supplementary Table S1. PRS variants and imputation in RS and NELSON.

| PRS | rsID | Chr | Pos | Discovery GWAS EA | NEA | EAF | Beta | *P*-value | Imputation quality (r-squared) RS-I | RS-II | RS-III | NELSON GenEx-I | NELSON GenEx-II |
|---|---|---|---|---|---|---|---|---|---|---|---|---|---|
| **PRS-McKay** | rs6695572 | 1 | 77945635 | A | G | 0.17 | 0.08 | 1.59E-07 | NA | NA | NA | NA | NA |
| | rs78062588* | 1 | 154566225 | C | T | 0.06 | -0.12 | 4.60E-07 | 0.910 | 0.895 | 0.909 | 0.920 | 0.888 |
| | rs78663305* | 1 | 168497741 | G | A | 0.12 | -0.10 | 2.55E-07 | 0.965 | 0.964 | 0.964 | 0.973 | 0.810 |
| | rs79368540* | 2 | 45189737 | T | C | 0.15 | 0.09 | 6.13E-07 | 0.923 | 0.913 | 0.915 | 0.933 | 0.877 |
| | rs11692700* | 2 | 67510377 | C | T | 0.03 | 0.18 | 6.44E-07 | 0.822 | 0.783 | 0.798 | 0.825 | 0.759 |
| | rs75442551 | 2 | 67915740 | A | C | 0.04 | 0.20 | 2.48E-07 | 0.671 | 0.700 | 0.730 | 0.720 | 0.793 |
| | rs1866631* | 2 | 174075761 | G | A | 0.40 | -0.06 | 6.97E-07 | 0.979 | 0.979 | 0.976 | 0.965 | 0.916 |
| | rs185666783* | 4 | 67833774 | G | C | 0.29 | -0.08 | 9.20E-08 | 0.991 | 0.990 | 0.991 | 0.975 | 0.925 |
| | rs77545136* | 5 | 1000080 | T | A | 0.03 | 0.20 | 8.29E-07 | 0.871 | 0.848 | 0.853 | 0.907 | 0.893 |
| | rs13156167 | 5 | 1275857 | C | T | 0.13 | 0.13 | 1.02E-10 | 0.546 | 0.563 | 0.663 | 0.691 | 0.825 |
| | rs7726159* | 5 | 1282319 | A | C | 0.35 | 0.09 | 1.93E-11 | 0.749 | 0.765 | 0.796 | 0.801 | 0.986 |
| | rs2736109* | 5 | 1296759 | T | C | 0.40 | 0.12 | 2.98E-19 | 0.725 | 0.724 | 0.771 | 0.775 | 0.998 |
| | rs2735947* | 5 | 1299392 | A | G | 0.14 | -0.12 | 4.32E-11 | 0.738 | 0.724 | 0.814 | 0.817 | 0.890 |
| | rs2853668* | 5 | 1300025 | T | G | 0.24 | 0.08 | 9.36E-09 | 0.997 | 0.997 | 0.997 | 0.987 | 0.909 |
| | rs112401627 | 5 | 1300269 | A | G | 0.03 | 0.27 | 3.03E-10 | 0.664 | 0.679 | 0.693 | 0.727 | 0.816 |
| | rs397640* | 5 | 1334729 | T | C | 0.09 | -0.12 | 5.58E-09 | 0.837 | 0.808 | 0.875 | 0.880 | 0.914 |
| | rs27066 | 5 | 1359133 | T | C | 0.13 | 0.11 | 6.63E-09 | 0.735 | 0.737 | 0.766 | 0.687 | 0.719 |
| | rs10050865* | 5 | 1360487 | T | C | 0.32 | -0.07 | 6.29E-08 | 0.855 | 0.857 | 0.879 | 0.880 | 0.835 |
| | rs9295661* | 6 | 25450026 | C | A | 0.06 | 0.12 | 7.83E-07 | 0.975 | 0.983 | 0.979 | 0.915 | 0.870 |
| | rs62396185* | 6 | 26180634 | C | G | 0.28 | -0.08 | 1.53E-09 | 0.957 | 0.960 | 0.957 | 0.952 | 0.972 |
| | rs4711112* | 6 | 26470862 | A | G | 0.21 | -0.07 | 9.61E-07 | 0.995 | 0.996 | 0.997 | 0.974 | 0.975 |
| | rs116718612 | 6 | 29603582 | G | C | 0.32 | 0.08 | 7.42E-09 | 0.230 | 0.234 | 0.225 | 0.951 | 0.949 |
| | rs141747817* | 6 | 29660940 | G | A | 0.37 | 0.06 | 6.27E-07 | 0.768 | 0.780 | 0.768 | 0.997 | 0.999 |
| | rs116104192 | 6 | 30338247 | A | G | 0.37 | 0.06 | 3.16E-07 | 0.000 | 0.000 | 0.000 | 0.994 | 0.997 |
| | rs116729111 | 6 | 30999065 | C | T | 0.38 | -0.07 | 4.16E-08 | 0.180 | 0.179 | 0.182 | 0.996 | 0.998 |
| | rs114096584 | 6 | 31063328 | T | C | 0.33 | 0.07 | 2.10E-08 | 0.671 | 0.670 | 0.671 | 0.999 | 0.999 |
| | rs114941286* | 6 | 31109882 | T | G | 0.48 | 0.06 | 5.43E-07 | 0.869 | 0.858 | 0.874 | 0.928 | 0.999 |
| | rs115114531 | 6 | 31335330 | C | G | 0.29 | 0.07 | 2.53E-08 | 0.516 | 0.470 | 0.515 | 1.000 | 0.999 |
| | rs111677893 | 6 | 31347069 | T | C | 0.26 | 0.08 | 1.33E-08 | 0.476 | 0.427 | 0.471 | 0.997 | 0.998 |
| | rs115441327 | 6 | 31613739 | C | T | 0.25 | 0.08 | 5.41E-09 | 0.694 | 0.681 | 0.696 | 0.999 | 0.987 |
| | rs115350531 | 6 | 32195935 | C | T | 0.35 | -0.06 | 3.81E-07 | 0.106 | 0.103 | 0.108 | 0.988 | 0.998 |
| | rs145864686 | 6 | 32340070 | T | C | 0.35 | 0.06 | 9.65E-07 | 0.018 | 0.017 | 0.018 | 0.956 | 0.965 |
| | rs9270383 | 6 | 32558298 | T | G | 0.26 | 0.08 | 1.17E-07 | 0.005 | 0.005 | 0.005 | 0.150 | 0.205 |
| | rs2647074 | 6 | 32574360 | T | C | 0.30 | -0.07 | 5.24E-07 | 0.393 | 0.384 | 0.396 | 0.842 | 0.988 |
| | rs115625939 | 6 | 32583584 | G | A | 0.13 | -0.11 | 6.21E-08 | 0.401 | 0.403 | 0.408 | 0.909 | 0.945 |
| | rs190788477 | 6 | 32597561 | A | G | 0.30 | 0.07 | 1.17E-07 | 0.015 | 0.015 | 0.015 | 0.608 | 0.891 |
| | rs77226909 | 6 | 32603854 | T | C | 0.21 | 0.09 | 2.07E-08 | 0.002 | 0.002 | 0.002 | 0.303 | 0.821 |
| | rs114793377 | 6 | 32741487 | G | C | 0.36 | 0.06 | 9.97E-08 | 0.428 | 0.429 | 0.432 | 0.985 | 0.992 |
| | rs115497191* | 6 | 32904039 | A | T | 0.07 | 0.11 | 3.23E-07 | 0.866 | 0.857 | 0.841 | 0.995 | 0.995 |
| | rs117534741* | 6 | 72384541 | A | G | 0.02 | 0.21 | 4.68E-07 | 0.895 | 0.897 | 0.884 | 0.887 | 0.995 |
| | rs4710145* | 6 | 167362976 | G | A | 0.47 | 0.06 | 1.21E-07 | 0.965 | 0.962 | 0.964 | 0.963 | 0.961 |

| | | | | | | | | | | | | | |
|---|---|---|---|---|---|---|---|---|---|---|---|---|---|
| | rs880991* | 8 | 27276705 | A | G | 0.05 | -0.13 | 7.24E-07 | 0.969 | 0.964 | 0.971 | 0.958 | 0.916 |
| | rs2565064* | 8 | 27327841 | C | G | 0.29 | 0.07 | 4.58E-07 | 0.998 | 0.998 | 0.998 | 0.985 | 0.941 |
| | rs149467044* | 8 | 27411100 | G | T | 0.16 | -0.08 | 8.95E-07 | 0.940 | 0.940 | 0.937 | 0.928 | 0.910 |
| | rs62560775* | 9 | 22052068 | G | A | 0.10 | 0.10 | 6.02E-07 | 0.936 | 0.930 | 0.948 | 0.960 | 0.940 |
| | rs191205566* | 9 | 102587233 | T | C | 0.02 | 0.34 | 1.17E-07 | 0.745 | 0.732 | 0.745 | 0.771 | 0.962 |
| | rs62621207* | 10 | 102672248 | T | A | 0.05 | 0.15 | 5.85E-07 | 0.844 | 0.817 | 0.852 | 0.834 | NA |
| | rs78853063* | 11 | 57250026 | T | C | 0.08 | -0.12 | 4.65E-07 | 0.940 | 0.936 | 0.942 | 0.946 | 0.998 |
| | rs78334599 | 11 | 115998756 | A | G | 0.04 | -0.16 | 7.93E-07 | 0.698 | 0.708 | 0.695 | 0.713 | 0.673 |
| | rs11216819* | 11 | 118084980 | G | A | 0.46 | 0.06 | 1.78E-07 | 0.990 | 0.991 | 0.991 | 0.967 | 0.917 |
| | rs7309927* | 12 | 1000185 | G | A | 0.16 | -0.08 | 7.86E-07 | 0.990 | 0.993 | 0.992 | 0.987 | 0.974 |
| | rs73351723* | 12 | 58831070 | A | G | 0.13 | 0.09 | 3.81E-07 | 0.953 | 0.946 | 0.955 | 0.960 | 0.998 |
| | rs56404467* | 13 | 32839990 | A | G | 0.02 | 0.23 | 1.11E-07 | 0.798 | 0.795 | 0.782 | 0.839 | 0.776 |
| | rs715693* | 15 | 47488977 | T | C | 0.40 | 0.06 | 9.53E-08 | 0.999 | 0.999 | 0.999 | 0.997 | 0.996 |
| | rs34895054* | 15 | 49285001 | C | G | 0.27 | -0.07 | 1.46E-07 | 0.950 | 0.946 | 0.949 | 0.949 | 0.794 |
| | rs2069520* | 15 | 75038486 | G | T | 0.03 | 0.17 | 6.74E-07 | 0.928 | 0.934 | 0.966 | 0.959 | 0.980 |
| | rs2869030 | 15 | 78711803 | G | T | 0.24 | -0.16 | 5.17E-28 | 0.714 | 0.702 | 0.719 | 0.683 | 0.690 |
| | rs4436747* | 15 | 78714008 | G | A | 0.44 | -0.08 | 2.87E-11 | 0.833 | 0.821 | 0.836 | 0.800 | 0.789 |
| | rs78385743* | 15 | 78750388 | T | A | 0.04 | -0.19 | 8.12E-09 | 0.728 | 0.716 | 0.739 | 0.733 | 0.825 |
| | rs16969892 | 15 | 78774737 | G | A | 0.03 | -0.21 | 6.23E-07 | 0.481 | 0.448 | 0.531 | 0.545 | 0.600 |
| | rs117131212* | 15 | 78819478 | G | A | 0.05 | 0.14 | 7.42E-07 | 0.794 | 0.781 | 0.796 | 0.779 | 0.722 |
| | rs141147481 | 15 | 78835239 | G | C | 0.02 | -0.27 | 8.53E-08 | 0.596 | 0.630 | 0.597 | 0.656 | 0.699 |
| | rs72740960* | 15 | 78851724 | G | A | 0.02 | 0.23 | 1.38E-07 | 0.783 | 0.749 | 0.752 | 0.716 | 1.000 |
| | rs116991229 | 15 | 78874555 | C | T | 0.02 | 0.37 | 9.54E-14 | 0.589 | 0.551 | 0.570 | 0.548 | 0.887 |
| | rs12441088* | 15 | 78928264 | G | T | 0.26 | -0.19 | 7.43E-42 | 0.899 | 0.893 | 0.899 | 0.888 | 0.911 |
| | rs3971828 | 15 | 79040042 | T | C | 0.28 | 0.10 | 3.11E-08 | 0.567 | 0.585 | 0.605 | 0.657 | 0.628 |
| | rs112827102 | 15 | 79058968 | C | T | 0.08 | -0.16 | 5.24E-10 | 0.458 | 0.471 | 0.485 | 0.599 | 0.715 |
| | rs28450923 | 15 | 79065557 | G | A | 0.13 | -0.13 | 7.76E-08 | 0.378 | 0.364 | 0.428 | 0.466 | 0.519 |
| | rs10163145* | 15 | 79075335 | A | G | 0.43 | 0.13 | 1.19E-25 | 0.756 | 0.763 | 0.847 | 0.911 | 0.935 |
| | rs146722791 | 15 | 79140191 | T | C | 0.08 | 0.15 | 1.73E-08 | 0.422 | 0.398 | 0.461 | 0.473 | 0.550 |
| | rs17181550* | 17 | 70299958 | G | T | 0.43 | -0.06 | 1.98E-07 | 1.000 | 1.000 | 1.000 | 0.999 | 0.872 |
| | rs66500423* | 19 | 41195170 | C | T | 0.30 | 0.07 | 2.23E-07 | 0.987 | 0.988 | 0.986 | 0.982 | 0.756 |
| | rs4803356* | 19 | 41207206 | G | C | 0.07 | -0.12 | 8.31E-07 | 0.944 | 0.952 | 0.953 | 0.966 | 0.868 |
| | rs145256949* | 19 | 41332357 | G | A | 0.28 | 0.07 | 3.95E-07 | 0.748 | 0.747 | 0.810 | 0.885 | 0.792 |
| | rs11878604 | 19 | 41333284 | C | T | 0.09 | -0.14 | 5.99E-09 | 0.558 | 0.554 | 0.618 | 0.922 | 0.730 |
| | rs4001921* | 19 | 41363301 | C | T | 0.25 | -0.08 | 1.09E-07 | 0.950 | 0.947 | 0.971 | 0.917 | 0.921 |
| **PRS-Byun** | rs34517439* | 1 | 78450517 | A | C | 0.10 | 0.15 | 5.20E-12 | 0.869 | 0.869 | 0.870 | 0.876 | 0.998 |
| | rs36108040* | 3 | 189335844 | G | A | 0.48 | -0.07 | 1.13E-07 | 0.928 | 0.975 | 0.971 | 0.983 | 0.999 |
| | rs13167280 | 5 | 1280477 | A | G | 0.14 | 0.14 | 8.79E-14 | 0.658 | 0.672 | 0.737 | 0.760 | 0.921 |
| | rs7725218* | 5 | 1282414 | A | G | 0.37 | 0.08 | 8.23E-10 | 0.774 | 0.787 | 0.812 | 0.809 | 0.998 |
| | rs2853677* | 5 | 1287194 | A | G | 0.55 | -0.15 | 3.37E-29 | 0.797 | 0.803 | 0.830 | 0.789 | 0.989 |
| | rs148297846* | 5 | 1298017 | A | G | 0.08 | 0.16 | 6.03E-11 | 0.726 | 0.724 | 0.786 | 0.810 | 0.954 |
| | rs2735947* | 5 | 1299392 | A | G | 0.13 | -0.13 | 3.36E-11 | 0.738 | 0.724 | 0.814 | 0.817 | 0.890 |
| | rs112401627 | 5 | 1300269 | A | G | 0.03 | 0.26 | 3.50E-11 | 0.664 | 0.679 | 0.693 | 0.727 | 0.816 |
| | rs2735845* | 5 | 1300584 | G | C | 0.21 | 0.12 | 1.70E-14 | 0.840 | 0.841 | 0.867 | 0.858 | 0.929 |
| | rs397640* | 5 | 1334729 | T | C | 0.10 | -0.13 | 1.80E-09 | 0.837 | 0.808 | 0.875 | 0.880 | 0.914 |

| | | | | | | | | | | | | |
|---|---|---|---|---|---|---|---|---|---|---|---|---|
| rs31487* | 5 | 1341101 | C | G | 0.42 | -0.16 | 7.55E-35 | 0.983 | 0.981 | 0.983 | 0.944 | 0.982 |
| rs56397275* | 5 | 1368956 | T | C | 0.26 | -0.09 | 2.76E-09 | 0.910 | 0.883 | 0.956 | 0.961 | 0.930 |
| rs12203592* | 6 | 396321 | T | C | 0.14 | 0.11 | 7.69E-09 | 0.998 | 0.998 | 0.998 | 0.987 | 0.988 |
| rs2316515* | 6 | 410848 | G | A | 0.58 | 0.07 | 3.21E-07 | 0.992 | 0.991 | 0.991 | 0.979 | 0.999 |
| rs72845208* | 6 | 27395664 | A | C | 0.07 | -0.13 | 1.71E-07 | 0.998 | 0.998 | 0.998 | 0.997 | 0.968 |
| rs35193884 | 6 | 31249611 | A | G | 0.07 | -0.13 | 8.82E-07 | 0.248 | 0.231 | 0.252 | 0.987 | 0.990 |
| rs34102154 | 6 | 32572106 | A | G | 0.16 | -0.11 | 1.72E-09 | 0.275 | 0.276 | 0.283 | 0.824 | 0.976 |
| rs17534632* | 6 | 109740101 | T | C | 0.20 | 0.10 | 3.82E-09 | 0.999 | 0.999 | 0.998 | 0.996 | 0.998 |
| rs398278* | 6 | 167388169 | G | A | 0.53 | -0.06 | 3.17E-07 | 0.986 | 0.985 | 0.985 | 0.961 | 0.966 |
| rs10265693* | 7 | 130720805 | G | A | 0.10 | 0.11 | 3.58E-07 | 0.998 | 0.998 | 0.998 | 0.987 | 0.986 |
| rs72477506* | 8 | 27394257 | T | C | 0.06 | -0.18 | 5.43E-11 | 0.894 | 0.904 | 0.884 | 0.885 | 0.863 |
| rs2640726* | 8 | 27405809 | C | T | 0.63 | 0.07 | 4.86E-07 | 0.904 | 0.912 | 0.903 | 0.894 | 0.885 |
| rs7823498* | 8 | 32403573 | T | C | 0.78 | -0.07 | 9.40E-07 | 0.990 | 0.992 | 0.990 | 0.981 | 1.000 |
| rs7850447* | 9 | 21763748 | A | G | 0.10 | 0.12 | 4.30E-08 | 0.991 | 0.988 | 0.985 | 0.990 | 0.929 |
| rs77560034* | 9 | 21964866 | C | G | 0.08 | 0.12 | 3.06E-07 | 0.909 | 0.902 | 0.911 | 0.912 | 0.932 |
| rs7902587* | 10 | 105694301 | T | C | 0.11 | 0.11 | 1.86E-07 | 0.973 | 0.970 | 0.973 | 0.983 | NA |
| rs11607355* | 11 | 118093547 | T | C | 0.50 | -0.06 | 7.74E-07 | 0.972 | 0.975 | 0.975 | 0.957 | 0.921 |
| rs10849602* | 12 | 1062479 | G | A | 0.52 | 0.07 | 6.87E-08 | 0.988 | 0.989 | 0.989 | 0.958 | 1.000 |
| rs118006060* | 12 | 2220237 | T | C | 0.03 | -0.19 | 5.89E-07 | 0.946 | 0.950 | 0.948 | 0.959 | 0.972 |
| rs11571818* | 13 | 32968810 | C | T | 0.01 | 0.45 | 3.50E-11 | 0.845 | 0.842 | 0.803 | 0.872 | 1.000 |
| rs2413932* | 15 | 49383481 | T | C | 0.73 | 0.09 | 1.70E-10 | 1.000 | 1.000 | 1.000 | 1.000 | 0.694 |
| rs72740960* | 15 | 78851724 | G | A | 0.02 | 0.23 | 8.27E-07 | 0.783 | 0.749 | 0.752 | 0.716 | 1.000 |
| rs55781567* | 15 | 78857986 | G | C | 0.38 | 0.22 | 1.89E-66 | 0.966 | 0.965 | 0.968 | 0.969 | 0.990 |
| rs2229961 | 15 | 78880752 | A | G | 0.02 | 0.30 | 5.06E-09 | 0.505 | 0.464 | 0.499 | 0.478 | 0.994 |
| rs12443170* | 15 | 78907736 | A | G | 0.13 | -0.19 | 4.32E-24 | 0.807 | 0.800 | 0.820 | 0.821 | 0.925 |
| rs117832812* | 15 | 78912472 | A | C | 0.07 | -0.20 | 7.79E-15 | 0.726 | 0.727 | 0.784 | 0.771 | 0.951 |
| rs12905116 | 15 | 79042734 | A | T | 0.74 | 0.09 | 2.19E-10 | 0.610 | 0.629 | 0.649 | 0.717 | 0.660 |
| rs3861180 | 15 | 79043393 | G | A | 0.94 | 0.15 | 1.12E-08 | 0.627 | 0.638 | 0.662 | 0.488 | 0.529 |
| rs4420501* | 15 | 79138478 | T | A | 0.46 | 0.09 | 5.91E-12 | 0.907 | 0.907 | 0.972 | 0.937 | 0.961 |
| rs75772022* | 16 | 15770416 | G | C | 0.02 | -0.26 | 1.76E-07 | 0.913 | 0.872 | 0.918 | 0.898 | 0.861 |
| rs56113850* | 19 | 41353107 | C | T | 0.58 | 0.10 | 1.54E-13 | 0.771 | 0.776 | 0.828 | 0.893 | 0.880 |
| rs61541144* | 20 | 62527305 | A | G | 0.08 | -0.12 | 6.79E-07 | 0.860 | 0.859 | 0.861 | 0.887 | 0.999 |
| rs17002246* | 21 | 19340411 | G | A | 0.03 | -0.20 | 9.15E-07 | 0.959 | 0.908 | 0.949 | 0.976 | 0.979 |

RsID: reference SNP cluster ID, Chr: chromosome, Pos: position (GRCh37/hg19), EA: effect allele, NEA: non-effect allele, EAF: effect allele frequency.

* Variant included in the final PRS.

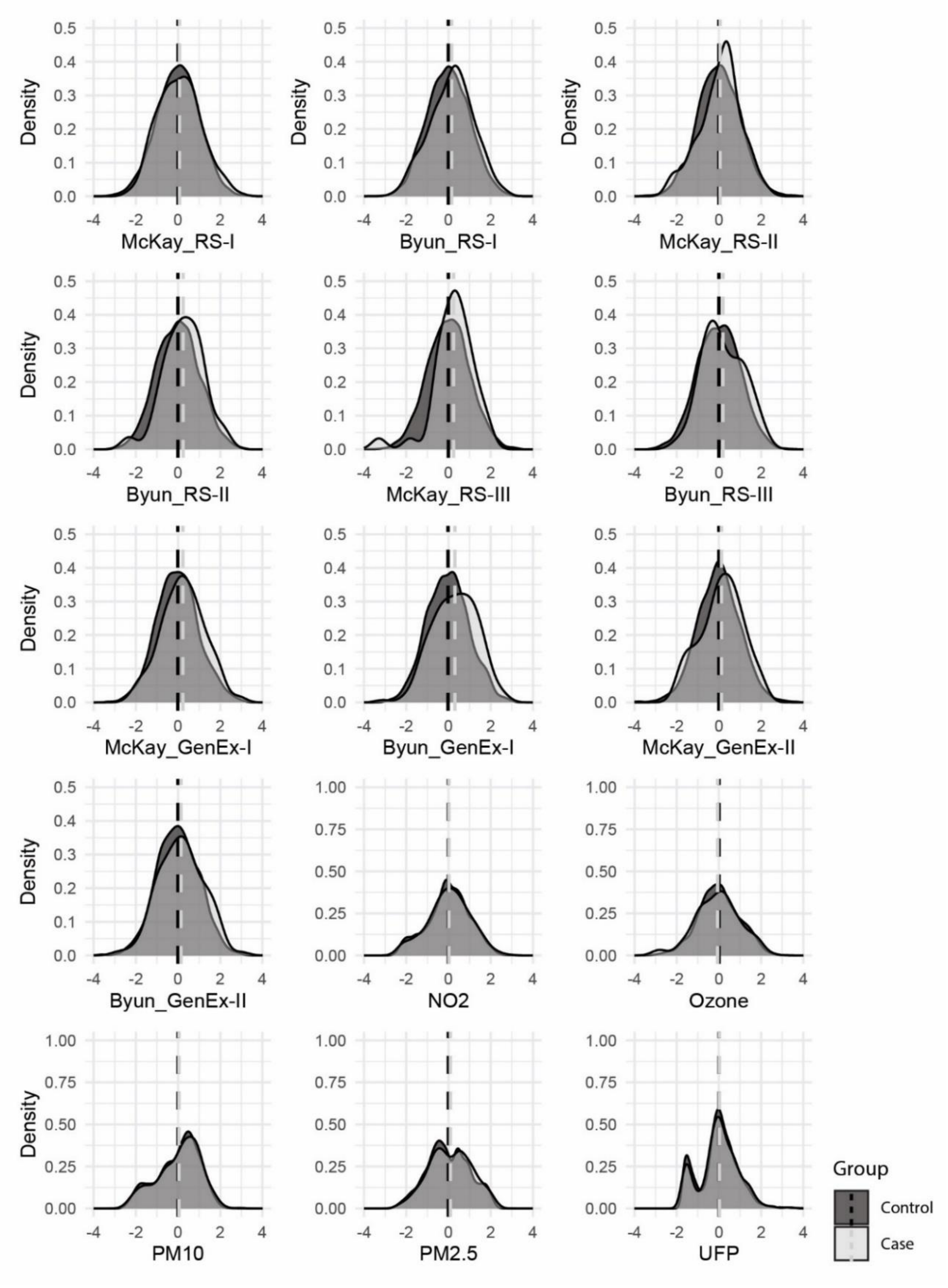

**Supplementary Figure S1. Distributions of exposures between lung cancer cases and controls.**

$NO_2$: nitrogen dioxide, $O_3$: ozone, $PM_{10}$: particulate matter <10 μm in diameter, $PM_{2.5}$: particulate matter <2.5 μm in diameter, UFP: ultra-fine particles; <0.1 μm in diameter.

**Supplementary Table S2. RS and NELSON subcohort demographics and clinical characteristics.**

| | RS (N = 14,926) | Genotyped RS (N = 11,493) | RS-I* (N = 6,291) | RS-II* (N = 2,154) | RS-III* (N = 3,048) | NELSON (N=7,364) | NELSON GenEx-I (N=2,896) | NELSON GenEx-II (N=2,308) | NELSON Ex (N=2,160) |
|---|---|---|---|---|---|---|---|---|---|
| **Demographics** | | | | | | | | | |
| Female sex, N (%) | 8,823 (59) | 6,670 (58) | 3,782 (60) | 1,171 (54) | 1,717 (56) | 1,196 (16) | 32 (1) | 495 (21) | 669 (31) |
| Age at cohort entry, years, median (IQR) | 63 (58-73) | 62 (58-71) | 68 (62-75) | 61 (58-69) | 56 (52-60) | 58 (54-62) | 58 (55-62) | 58 (54-63) | 57 (54-62) |
| **Smoking Status at Cohort Entry**, n (%) | | | | | | | | | |
| - Current | 3,116 (21) | 2,382 (21) | 1,282 (20) | 416 (19) | 684 (22) | 4,097 (56) | 1626 (56) | 1,249 (54) | 1,222 (57) |
| - Former | 6,159 (41) | 4,914 (43) | 2,445 (39) | 1,078 (50) | 1,391 (46) | 3,267 (44) | 1270 (44) | 1,059 (46) | 938 (43) |
| - Never | 5,070 (34) | 3,882 (34) | 2,259 (36) | 657 (31) | 966 (32) | - | - | - | - |
| - Missing | 581 (4) | 315 (3) | 305 (5) | 3 (0) | 7 (0) | 0 (0) | 0 (0) | 0 (0) | 0 (0) |
| Pack-years current smokers, median (IQR) | 29 (17-44) | 30 (17-44) | 30 (18-44) | 31 (16-44) | 29 (16-41) | 38 (28-50) | 38 (28-46) | 38 (30-50) | 38 (30-50) |
| - Missing, n (% of current smokers) | 189 (6) | 128 (5) | 40 (3) | 30 (7) | 58 (9) | 0 (0) | 0 (0) | 0 (0) | 0 (0) |
| Pack-years former smokers, median (IQR) | 16 (5-33) | 16 (5-32) | 18 (6-36) | 15 (5-32) | 13 (5-26) | 38 (30-50) | 38 (30-46) | 39 (30-50) | 38 (30-53) |
| - Missing, n (% of former smokers) | 532 (9) | 415 (8) | 226 (9) | 83 (8) | 106 (8) | 0 (0) | 0 (0) | 0 (0) | 0 (0) |
| **Environmental Exposure**, mean ± SD | | | | | | | | | |
| - $NO_2$, ug/$m_3$ | - | - | - | - | - | 27 ± 7 | 24 ± 7 | 29 ± 5 | 29 ± 6 |
| - $O_3$, ug/$m_3$ | - | - | - | - | - | 56 ± 4 | 57 ± 5 | 55 ± 4 | 55 ± 4 |
| - $PM_{10}$, ug/$m_3$ | - | - | - | - | - | 29 ± 3 | 28 ± 3 | 30 ± 2 | 30 ± 2 |
| - $PM_{2.5}$, ug/$m_3$ | - | - | - | - | - | 16 ± 1 | 16 ± 2 | 16 ± 1 | 16 ± 1 |
| - UFP, particles/$m_3$ | - | - | - | - | - | 11,477 ± 3,628 | 10,313 ± 3,158 | 12,416 ± 4,139 | 12,034 ± 3,185 |
| **Lung Cancer Diagnoses** | | | | | | | | | |
| Lung cancer cases, n (%) | 449 (3) | 356 (3) | 260 (4) | 60 (3) | 36 (1) | 469 (6) | 166 (6) | 115 (5) | 188 (9) |
| Age-at-diagnosis, years, median (IQR) | 76 (69-81) | 76 (69-80) | 77 (71-81) | 73 (69-77) | 61 (56-66) | 70 (65-74) | 70 (66-75) | 71 (66-75) | 68 (64-74) |
| Histological subtype, n (% of lung cancer) | | | | | | | | | |
| - NSCLC | 298 (66) | 242 (68) | 174 (67) | 41 (68) | 27 (75) | 363 (77) | 128 (77) | 87 (76) | 148 (79) |
| - ADN | 84 (19) | 65 (18) | 44 (17) | 10 (17) | 11 (31) | 244 (52) | 80 (48) | 62 (54) | 102 (54) |
| - SQM | 101 (23) | 84 (24) | 64 (25) | 13 (22) | 7 (19) | 91 (19) | 42 (25) | 19 (17) | 30 (16) |
| - NSCLC-NOS | 113 (25) | 93 (26) | 66 (25) | 18 (30) | 9 (25) | 28 (6) | 6 (4) | 6 (5) | 16 (9) |
| - SCLC | 59 (13) | 47 (13) | 31 (12) | 12 (20) | 4 (11) | 62 (13) | 19 (11) | 20 (17) | 23 (12) |
| - Other/NOS/unknown | 92 (21) | 67 (19) | 55 (21) | 7 (12) | 5 (14) | 44 (9) | 19 (11) | 8 (7) | 17 (9) |
| **Screening Outcomes** | | | | | | | | | |
| Positive/indeterminate screening result, n | - | - | - | - | - | 2,214 (30) | 888 (31) | 679 (30) | 647 (30) |
| - Missing, n (%) | - | - | - | - | - | 263 (4) | 1 (0) | 2 (0) | 260 (12) |
| Lung cancer mortality, n (%) | - | - | - | - | - | 226 (3) | 85 (3) | 52 (2) | 89 (4) |
| All-cause mortality, n (%) | - | - | - | - | - | 1,214 (17) | 461 (16) | 359 (16) | 394 (18) |

Percentages are calculated as % of total unless specified otherwise. IQR: interquartile range, SD: standard deviation, $NO_2$: nitrogen dioxide, $O_3$: ozone, $PM_{10}$: particulate matter <10 μm in diameter, $PM_{2.5}$: particulate matter <2.5 μm in diameter, UFP: ultra-fine particles; <0.1 μm in diameter, ADN: adenocarcinoma, SQM: squamous cell carcinoma, SCLC: small cell lung cancer, NSCLC: non-small cell lung cancer, NOS: not otherwise specified.

*Data represent the genotyped subset of the RS cohort.

**Supplementary Table S3. Subcohort characteristics split by case-control status.**

| | RS Controls* (N = 11,137) | RS Cases* (N = 356) | NELSON Controls (N=6,895) | NELSON Cases (N=469) |
|---|---|---|---|---|
| **Demographics** | | | | |
| Female sex, n (%) | 6,532 (59) | 138 (39) | 1,121 (16) | 75 (16) |
| Age at cohort entry, years, median (IQR) | 62 (58-71) | 65 (60-70) | 58 (54-62) | 60 (56-65) |
| **Smoking Status at Cohort Entry**, n (%) | | | | |
| - Current | 2,194 (20) | 188 (53) | 3,797 (55) | 300 (64) |
| - Former | 4,794 (43) | 120 (34) | 3,098 (45) | 169 (36) |
| - Never | 3,849 (35) | 33 (9) | - | - |
| - Missing | 300 (3) | 15 (4) | 0 (0) | 0 (0) |
| Pack-years current smokers, median (IQR) | 29 (17-43) | 37 (25-50) | 38 (28-50) | 44 (34-55) |
| - Missing, n (% of current smokers) | 125 (6) | 3 (2) | 0 (0) | 0 (0) |
| Pack-years former smokers, median (IQR) | 15 (5-31) | 37 (23-58) | 38 (30-50) | 44 (34-60) |
| - Missing, n (% of former smokers) | 407 (9) | 8 (7) | 0 (0) | 0 (0) |
| **Environmental Exposure**, mean ± SD | | | | |
| - $NO_2$, ug/$m_3$ | - | - | 27 ± 7 | 27 ± 7 |
| - $O_3$, ug/$m_3$ | - | - | 56 ± 4 | 56 ± 5 |
| - $PM_{10}$, ug/$m_3$ | - | - | 29 ± 3 | 29 ± 3 |
| - $PM_{2.5}$, ug/$m_3$ | - | - | 16 ± 1 | 16 ± 1 |
| - UFP, particles/$m_3$ | - | - | 11,476 ± 3,671 | 11,496 ± 2,937 |
| **Screening Outcomes** | | | | |
| Positive/indeterminate screening result, n (%) | - | - | 1,884 (27) | 330 (70) |
| - Missing, n (%) | - | - | 231 (3) | 32 (7) |
| Lung cancer mortality, n (%) | - | - | 7 (0) | 219 (47) |
| All-cause mortality, n (%) | - | - | 929 (14) | 285 (61) |

Percentages are calculated as % of total unless specified otherwise. IQR: interquartile range, SD: standard deviation, $NO_2$: nitrogen dioxide, $O_3$: ozone, $PM_{10}$: particulate matter <10 µm in diameter, $PM_{2.5}$: particulate matter <2.5 µm in diameter, UFP: ultra-fine particles; <0.1 µm in diameter.

* Data represent the genotyped subset of the RS cohort.

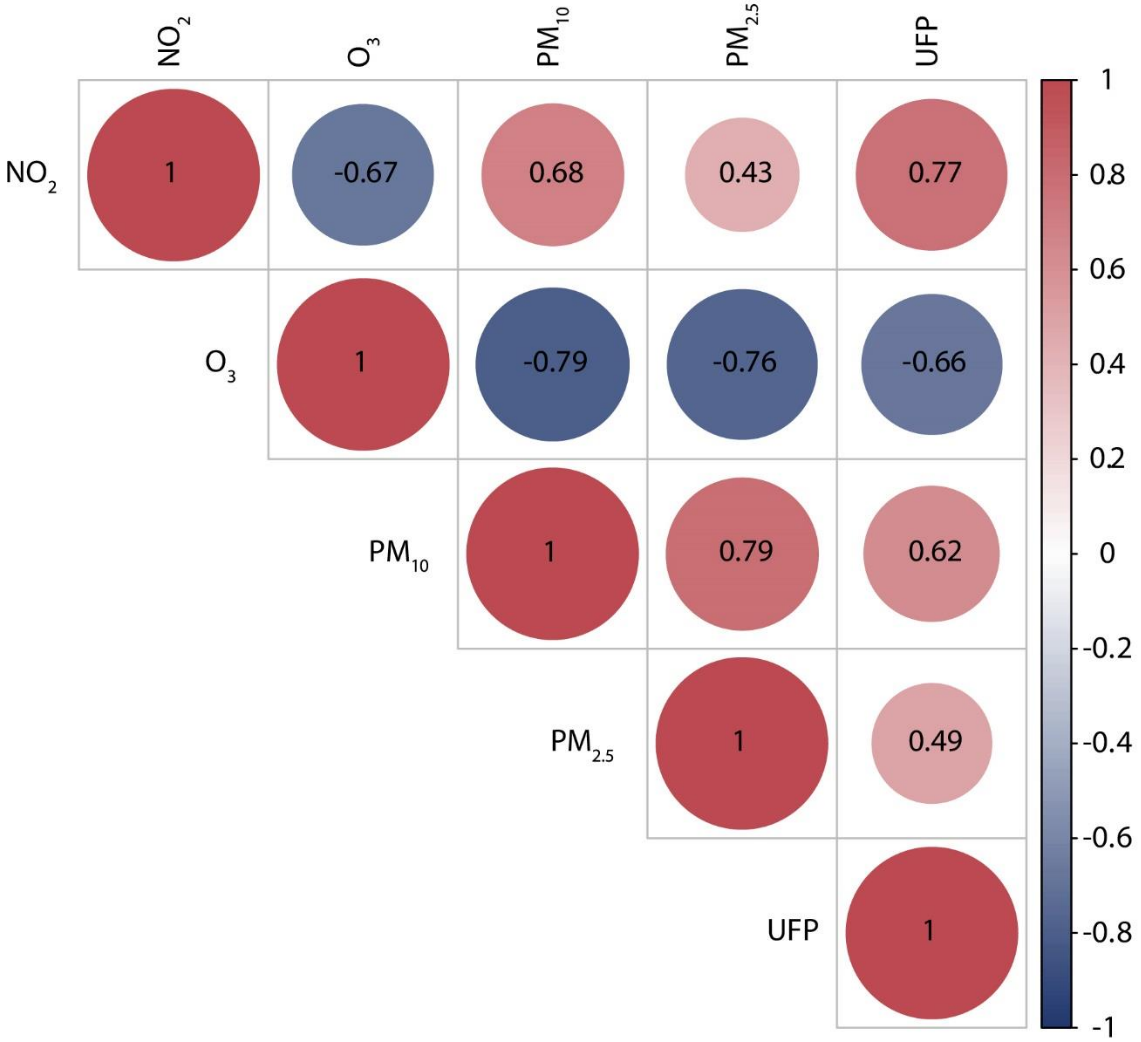

**Supplementary Figure S2. Pearson correlations (r) between ambient air pollution constituents in NELSON.**

$NO_2$: nitrogen dioxide, $O_3$: ozone, $PM_{10}$: particulate matter <10 µm in diameter, $PM_{2.5}$: particulate matter <2.5 µm in diameter, UFP: ultra-fine particles; <0.1 µm in diameter.

| Cohort | Strata | Category | Cases | Controls | PRS−McKay OR [95%CI] | PRS−Byun OR [95%CI] |
|---|---|---|---|---|---|---|
| RS | Sex | Male | 218 | 4605 | 1.07 [0.93−1.22] | 1.08 [0.94−1.24] |
| | | Female | 138 | 6532 | 1.14 [0.96−1.35] | 1.43 [1.21−1.69] |
| | Age−at−diagnosis | Below 75 | 162 | 11137 | 1.23 [1.06−1.44] | 1.30 [1.12−1.52] |
| | | Over 75 | 192 | 8125 | 0.99 [0.86−1.15] | 1.11 [0.96−1.28] |
| | Smoking status | Current | 188 | 2194 | 1.13 [0.97−1.32] | 1.27 [1.09−1.48] |
| | | Former | 120 | 4794 | 1.11 [0.92−1.33] | 1.14 [0.95−1.37] |
| | | Never | 32 | 2884 | 0.95 [0.67−1.33] | 1.21 [0.85−1.71] |
| | Lung cancer subtype | ADN | 65 | 11137 | 0.91 [0.72−1.17] | 1.33 [1.04−1.70] |
| | | SQM | 84 | 11137 | 1.07 [0.87−1.33] | 1.14 [0.92−1.41] |
| | | SCLC | 47 | 11137 | 1.18 [0.88−1.57] | 1.04 [0.78−1.38] |
| | | NSCLC-NOS | 93 | 11137 | 1.06 [0.86−1.30] | 1.13 [0.92−1.38] |
| NELSON | Sex | Male | 257 | 4420 | 1.26 [1.11−1.43] | 1.28 [1.13−1.46] |
| | | Female | 22 | 473 | 0.77 [0.51−1.18] | 1.15 [0.75−1.74] |
| | Age−at−diagnosis | Below 75 | 215 | 4923 | 1.15 [1.01−1.32] | 1.25 [1.09−1.43] |
| | | Over 75 | 66 | 4923 | 1.45 [1.14−1.84] | 1.39 [1.09−1.77] |
| | Smoking status | Current | 187 | 2688 | 1.18 [1.02−1.37] | 1.23 [1.06−1.42] |
| | | Former | 94 | 2235 | 1.31 [1.06−1.60] | 1.39 [1.12−1.72] |
| | Lung cancer subtype | ADN | 142 | 4923 | 1.15 [0.98−1.36] | 1.27 [1.07−1.50] |
| | | SQM | 61 | 4923 | 1.21 [0.94−1.55] | 1.20 [0.93−1.55] |
| | | SCLC | 39 | 4923 | 1.16 [0.85−1.59] | 1.34 [0.97−1.83] |
| | | NSCLC-NOS | 12 | 4923 | 2.62 [1.47−4.70] | 1.35 [0.77−2.39] |

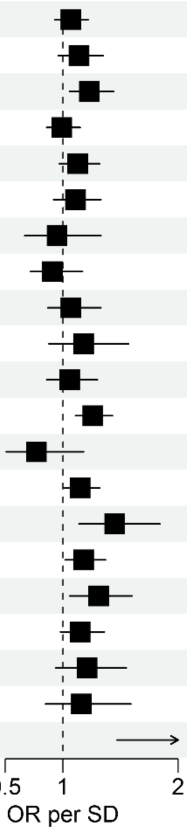

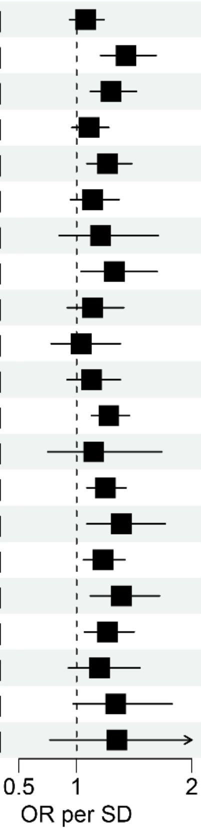

**Supplementary Figure S3. Stratified analyses of continuous PRSs in RS and NELSON.**

Generalized linear univariable model analyzing the PRSs, with binomial outcome: lung cancer yes/no. Fixed effects meta-analyses were performed on RS-I, RS-II, and RS-III, and NELSON GenEx-I, and GenEx-II. The results for the female stratum of the NELSON cohort correspond to only the NELSON GenEx-II subcohort. For smoking status ‘current’ and age-at-diagnosis ‘over 75’, the results for RS represent a meta-analysis of RS-I and RS-II. The OR corresponds to the increase in odds of lung cancer per standard deviation increase in the PRS. OR: odds ratio, SD: standard deviation, ADN: adenocarcinoma, SQM: squamous cell carcinoma, SCLC: small cell lung cancer, NSCLC: non-small cell lung cancer.

### Supplementary Table S4. Subcohort PRS results of main association analysis.

| Cohort | PRS | Cases | Controls | OR per SD [95% CI] | *P*-value | AUC [95% CI] |
|---|---|---|---|---|---|---|
| RS-I | McKay | 260 | 6031 | 1.07 [0.95-1.21] | 2.68E-01 | 0.513 [0.476-0.549] |
| RS-I | Byun | 260 | 6031 | 1.17 [1.03-1.32] | 1.48E-02 | 0.547 [0.511-0.583] |
| RS-II | McKay | 60 | 2094 | 1.06 [0.82-1.36] | 6.77E-01 | 0.527 [0.453-0.600] |
| RS-II | Byun | 60 | 2094 | 1.28 [0.99-1.66] | 5.63E-02 | 0.580 [0.510-0.649] |
| RS-III | McKay | 36 | 3012 | 1.28 [0.92-1.78] | 1.46E-01 | 0.593 [0.509-0.677] |
| RS-III | Byun | 36 | 3012 | 1.21 [0.87-1.68] | 2.59E-01 | 0.451 [0.357-0.545] |
| Meta-RS | McKay | 356 | 11137 | 1.09 [0.98-1.21] | 1.13E-01 | 0.523 [0.492-0.554] |
| Meta-RS | Byun | 356 | 11137 | 1.19 [1.07-1.32] | 1.25E-03 | 0.541 [0.510-0.572] |
| NELSON GenEx-I | McKay | 166 | 2738 | 1.28 [1.09-1.49] | 2.04E-03 | 0.572 [0.526-0.617] |
| NELSON GenEx-I | Byun | 166 | 2738 | 1.37 [1.17-1.60] | 7.95E-05 | 0.590 [0.544-0.637] |
| NELSON GenEx-II | McKay | 116 | 2201 | 1.14 [0.95-1.38] | 1.66E-01 | 0.545 [0.489-0.601] |
| NELSON GenEx-II | Byun | 116 | 2201 | 1.16 [0.96-1.39] | 1.30E-01 | 0.539 [0.483-0.594] |
| Meta-NELSON | McKay | 282 | 4939 | 1.22 [1.08-1.37] | 1.13E-03 | 0.560 [0.525-0.596] |
| Meta-NELSON | Byun | 282 | 4939 | 1.28 [1.13-1.44] | 6.41E-05 | 0.569 [0.533-0.604] |

Generalized univariable linear model analyzing the PRSs, with binomial outcome: lung cancer yes/no. The meta-RS and meta-NELSON analyses represent a fixed effects meta-analysis on RS-I, RS-II, and RS-III, and on NELSON GenEx-I, and GenEx-II. The OR corresponds to the increase in odds of lung cancer per standard deviation increase in the PRS. OR: odds ratio, SD: standard deviation, AUC: area under the curve.

**Supplementary Table S5. Sensitivity analysis for incident cases in RS.**

| Cohort | PRS | Cases | Controls | OR per SD [95% CI] | *P*-value | AUC [95% CI] |
|---|---|---|---|---|---|---|
| RS-I | McKay | 253 | 6031 | 1.08 [0.95-1.23] | 2.24E-01 | 0.515 [0.478-0.552] |
| RS-I | Byun | 253 | 6031 | 1.17 [1.03-1.33] | 1.35E-02 | 0.548 [0.511-0.585] |
| RS-II | McKay | 58 | 2094 | 1.05 [0.81-1.36] | 7.14E-01 | 0.525 [0.449-0.600] |
| RS-II | Byun | 58 | 2094 | 1.28 [0.99-1.67] | 6.04E-02 | 0.579 [0.508-0.651] |
| RS-III | McKay | 28 | 3012 | 1.14 [0.79-1.66] | 4.78E-01 | 0.555 [0.453-0.657] |
| RS-III | Byun | 28 | 3012 | 1.01 [0.70-1.46] | 9.61E-01 | 0.510 [0.403-0.617] |
| Meta-RS | McKay | 339 | 11137 | 1.08 [0.97-1.20] | 1.59E-01 | 0.520 [0.489-0.552] |
| Meta-RS | Byun | 339 | 11137 | 1.18 [1.05-1.31] | 3.47E-03 | 0.550 [0.518-0.582] |

Generalized univariable linear model analyzing the PRSs, with binomial outcome: lung cancer yes/no. The meta-RS analyses represent a fixed effects meta-analysis on RS-I, RS-II, and RS-III. The OR corresponds to the increase in odds of lung cancer per standard deviation increase in the PRS. OR: odds ratio, SD: standard deviation, AUC: area under the curve.

### Supplementary Table S6. Sensitivity analysis for histologically confirmed cases in RS.

| Cohort | PRS | Cases | Controls | OR per SD [95% CI] | *P*-value | AUC [95% CI] |
|---|---|---|---|---|---|---|
| RS-I | McKay | 220 | 6071 | 1.07 [0.94-1.23] | 3.19E-01 | 0.511 [0.471-0.552] |
| RS-I | Byun | 220 | 6071 | 1.15 [1.00-1.31] | 4.42E-02 | 0.542 [0.502-0.581] |
| RS-II | McKay | 56 | 2098 | 1.05 [0.80-1.37] | 7.30E-01 | 0.526 [0.451-0.600] |
| RS-II | Byun | 56 | 2098 | 1.28 [0.98-1.67] | 6.67E-02 | 0.578 [0.506-0.651] |
| RS-III | McKay | 35 | 3013 | 1.29 [0.92-1.80] | 1.34E-01 | 0.597 [0.511-0.683] |
| RS-III | Byun | 35 | 3013 | 1.23 [0.88-1.71] | 2.34E-01 | 0.447 [0.351-0.543] |
| Meta-RS | McKay | 311 | 11182 | 1.09 [0.97-1.22] | 1.36E-01 | 0.523 [0.490-0.556] |
| Meta-RS | Byun | 311 | 11182 | 1.18 [1.05-1.32] | 4.08E-03 | 0.536 [0.503-0.569] |

Generalized univariable linear model analyzing the PRSs, with binomial outcome: lung cancer yes/no. The OR corresponds to the increase in odds of lung cancer per standard deviation increase in the PRS and AAP exposures. The meta-RS analyses represent a fixed effects meta-analysis on RS-I, RS-II, and RS-III. OR: odds ratio, SD: standard deviation, AUC: area under the curve.

**Supplementary Table S7. Combined PRS performance across RS subcohorts.**

| Cohort | PRS | Cases | Controls | OR per SD [95% CI] | *P*-value | AUC [95% CI] |
|---|---|---|---|---|---|---|
| RS-I | McKay | 260 | 6031 | 1.07 [0.95-1.21] | 2.68E-01 | 0.513 [0.476-0.549] |
| RS-I | Byun | 260 | 6031 | 1.17 [1.03-1.32] | 1.48E-02 | 0.547 [0.511-0.583] |
| RS-I | Combined | 260 | 6031 | 1.06 [0.93-1.20] | 3.69E-01 | 0.516 [0.480-0.552] |
| RS-II | McKay | 60 | 2094 | 1.06 [0.82-1.36] | 6.77E-01 | 0.527 [0.453-0.600] |
| RS-II | Byun | 60 | 2094 | 1.28 [0.99-1.66] | 5.63E-02 | 0.580 [0.510-0.649] |
| RS-II | Combined | 60 | 2094 | 1.07 [0.82-1.38] | 6.30E-01 | 0.529 [0.458-0.600] |
| RS-III | McKay | 36 | 3012 | 1.28 [0.92-1.78] | 1.46E-01 | 0.593 [0.509-0.677] |
| RS-III | Byun | 36 | 3012 | 1.21 [0.87-1.68] | 2.59E-01 | 0.451 [0.357-0.545] |
| RS-III | Combined | 36 | 3012 | 1.41 [1.01-1.97] | 4.44E-02 | 0.612 [0.520-0.705] |
| Meta-RS | McKay | 356 | 11137 | 1.09 [0.98-1.21] | 1.13E-01 | 0.523 [0.492-0.554] |
| Meta-RS | Byun | 356 | 11137 | 1.19 [1.07-1.32] | 1.25E-03 | 0.541 [0.510-0.572] |
| Meta-RS | Combined | 356 | 11137 | 1.09 [0.98-1.21] | 1.09E-01 | 0.528 [0.497-0.559] |

Generalized univariable linear model analyzing the PRSs, with binomial outcome: lung cancer yes/no. The meta-RS analyses represent a fixed effects meta-analysis on RS-I, RS-II, and RS-III. The OR corresponds to the increase in odds of lung cancer per standard deviation increase in the PRS. OR: odds ratio, SD: standard deviation, AUC: area under the curve.

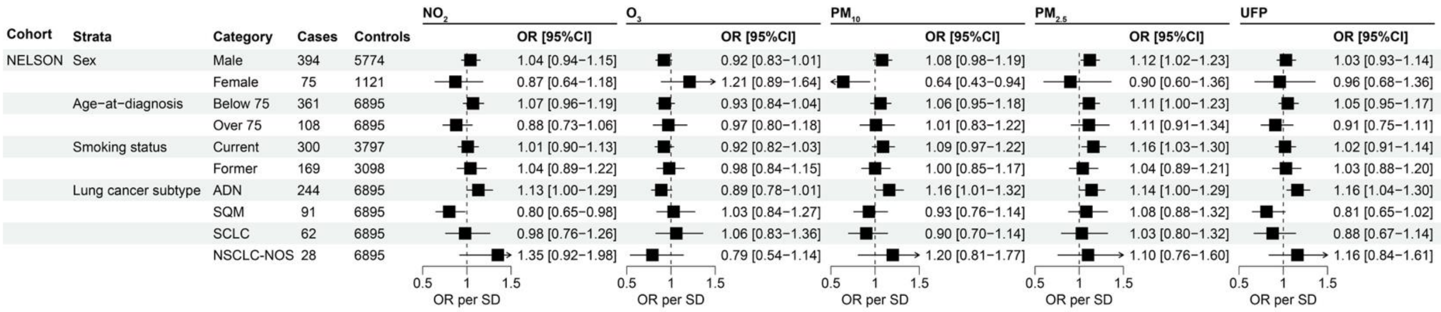

**Supplementary Figure S4. Stratified analysis of ambient air pollution constituents in NELSON.**

Generalized univariable linear model analyzing the AAP constituents, with binomial outcome: lung cancer yes/no. The OR corresponds to the increase in odds of lung cancer per standard deviation increase in the AAP exposures. OR: odds ratio, SD: standard deviation, $NO_2$: nitrogen dioxide, $O_3$: ozone, $PM_{10}$: particulate matter <10 μm in diameter, $PM_{2.5}$: particulate matter <2.5 μm in diameter, UFP: ultra-fine particles; <0.1 μm in diameter, ADN: adenocarcinoma, SQM: squamous cell carcinoma, SCLC: small cell lung cancer, NSCLC: non-small cell lung cancer.

**Supplementary Table S8. Analysis of pollutant exposures three years prior to randomization.**

| Cohort | Exposure | Cases | Controls | OR per SD [95% CI] | *P*-value | AUC [95% CI] |
|---|---|---|---|---|---|---|
| NELSON | $NO_2$ | 469 | 6895 | 1.02 [0.93-1.12] | 6.98E-01 | 0.497 [0.470-0.524] |
| NELSON | $O_3$ | 469 | 6895 | 0.96 [0.88-1.06] | 4.50E-01 | 0.492 [0.464-0.520] |
| NELSON | $PM_{10}$ | 469 | 6895 | 1.03 [0.94-1.13] | 5.22E-01 | 0.509 [0.482-0.536] |
| NELSON | $PM_{2.5}$ | 469 | 6895 | 1.02 [0.93-1.12] | 6.61E-01 | 0.504 [0.477-0.531] |

Generalized univariable linear model analyzing the AAP constituents, with binomial outcome: lung cancer yes/no. The OR corresponds to the increase in odds of lung cancer per standard deviation increase in the AAP exposure. OR: odds ratio, SD: standard deviation, AUC: area under the curve, $NO_2$: nitrogen dioxide, $O_3$: ozone, $PM_{10}$: particulate matter <10 µm in diameter, $PM_{2.5}$: particulate matter <2.5 µm in diameter.

| Exposure | Cohort | Decile | Cases | Controls | OR [95% CI] | *P*-value | AUC [95% CI] |
|---|---|---|---|---|---|---|---|
| Polygenic risk score | | | | | | | |
| PRS−McKay | RS | Bottom | 97 | 3352 | 1.08 [0.70−1.65] | 7.3e−01 | 0.520 [0.461−0.579] |
| | | Top | 108 | 3341 | 1.44 [0.98−2.13] | 6.6e−02 | 0.542 [0.486−0.599] |
| | NELSON | Bottom | 85 | 1476 | 1.04 [0.66−1.66] | 8.6e−01 | 0.525 [0.461−0.589] |
| | | Top | 97 | 1464 | 1.49 [0.98−2.26] | 6.3e−02 | 0.546 [0.486−0.607] |
| PRS−Byun | RS | Bottom | 96 | 3353 | 0.89 [0.57−1.40] | 6.2e−01 | 0.520 [0.461−0.579] |
| | | Top | 111 | 3338 | 1.33 [0.90−1.96] | 1.5e−01 | 0.533 [0.477−0.588] |
| | NELSON | Bottom | 63 | 1498 | 0.93 [0.54−1.59] | 7.8e−01 | 0.511 [0.438−0.584] |
| | | Top | 92 | 1469 | 2.40 [1.57−3.67] | 5.2e−05 | 0.605 [0.543−0.668] |
| Ambient air pollution | | | | | | | |
| $NO_2$ | NELSON | Bottom | 130 | 2079 | 1.06 [0.73−1.54] | 7.6e−01 | 0.507 [0.464−0.549] |
| | | Top | 134 | 2075 | 1.16 [0.81−1.67] | 4.2e−01 | 0.517 [0.475−0.559] |
| $O_3$ | NELSON | Bottom | 139 | 2070 | 1.47 [1.03−2.08] | 3.2e−02 | 0.545 [0.502−0.587] |
| | | Top | 128 | 2081 | 1.17 [0.81−1.69] | 4.1e−01 | 0.518 [0.475−0.561] |
| $PM_{10}$ | NELSON | Bottom | 142 | 2067 | 0.92 [0.64−1.33] | 6.6e−01 | 0.509 [0.469−0.549] |
| | | Top | 143 | 2066 | 0.94 [0.66−1.36] | 7.5e−01 | 0.506 [0.467−0.546] |
| $PM_{2.5}$ | NELSON | Bottom | 123 | 2086 | 1.00 [0.68−1.47] | 9.9e−01 | 0.500 [0.457−0.543] |
| | | Top | 132 | 2079 | 1.23 [0.86−1.77] | 2.6e−01 | 0.524 [0.481−0.566] |
| UFP | NELSON | Bottom | 145 | 2065 | 1.20 [0.85−1.70] | 3.0e−01 | 0.521 [0.480−0.562] |
| | | Top | 146 | 2064 | 1.22 [0.87−1.73] | 2.5e−01 | 0.523 [0.482−0.564] |

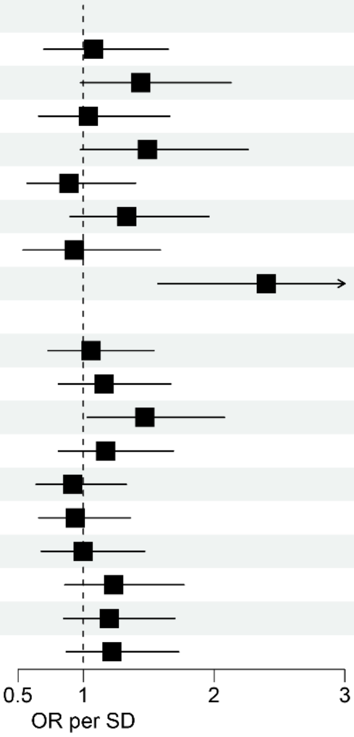

**Supplementary Figure S5. Top and bottom deciles of PRS and ambient air pollution constituents.**

Generalized univariable linear model analyzing top and bottom deciles compared to the center of the distribution (40th-60th percentile), with binomial outcome: lung cancer yes/no. For PRS analyses, fixed effects meta-analyses were performed on RS-I, RS-II, and RS-III, and NELSON GenEx-I, and GenEx-II. OR: odds ratio, SD: standard deviation, AUC: area under the curve, $NO_2$: nitrogen dioxide, $O_3$: ozone, $PM_{10}$: particulate matter <10 µm in diameter, $PM_{2.5}$: particulate matter <2.5 µm in diameter, UFP: ultra-fine particles; <0.1 µm in diameter.

**Supplementary Table S9. Association of PRSs and AAP constituents with indeterminate/false-positive CT results.**

| Cohort | Exposure | Cases | Controls | OR per SD [95% CI] | *P*-value |
|---|---|---|---|---|---|
| NELSON | PRS-McKay | 1378 | 3542 | 1.07 [1.01-1.14] | 2.93E-02 |
| NELSON | PRS-Byun | 1378 | 3542 | 1.01 [0.94-1.07] | 8.63E-01 |
| NELSON | $NO_2$ | 1884 | 4780 | 0.99 [0.94-1.04] | 7.22E-01 |
| NELSON | $O_3$ | 1884 | 4780 | 1.00 [0.95-1.05] | 9.95E-01 |
| NELSON | $PM_{10}$ | 1884 | 4780 | 1.04 [0.98-1.09] | 1.97E-01 |
| NELSON | $PM_{2.5}$ | 1884 | 4780 | 1.01 [0.95-1.06] | 8.54E-01 |
| NELSON | UFP | 1884 | 4780 | 1.01 [0.96-1.06] | 7.70E-01 |

Generalized univariable linear model analyzing the PRSs and AAP constituents, with binomial outcome: indeterminate/positive CT result in one of the four screening rounds, requiring follow-up imaging or clinical referral, restricted to participants without lung cancer after follow-up. The PRS analyses represent a fixed effects meta-analysis on NELSON GenEx-I, and GenEx-II. The OR corresponds to the increase in odds of lung cancer per standard deviation increase in the PRS and AAP exposures. LC: lung cancer, OR: odds ratio, SD: standard deviation, $NO_2$: nitrogen dioxide, $O_3$: ozone, $PM_{10}$: particulate matter <10 µm in diameter, $PM_{2.5}$: particulate matter <2.5 µm in diameter, UFP: ultra-fine particles; <0.1 µm in diameter.
